# Development and Clinical Evaluation of AlcoChange: a Digital Therapeutic for Patients with Alcohol-Related Liver Disease

**DOI:** 10.1101/2023.08.22.23293936

**Authors:** G Mehta, S Lin, A Nadar, PV Bharadwaj, R Kumar, A Balaji, S Macdonald, M Sheikh, MM Saeidinejad, SR Sharma, JJ King, RP Mookerjee, L McDonagh, SA Afuwape, K Moore, R Jalan

**Author notes:** **Corresponding author:** Gautam Mehta. **Conflict of Interest**. RJ is the founder of Cyberliver Ltd, the company that holds the intellectual property for AlcoChange. He is also the founder of Yaqrit Discovery and Hepyx Limited, and inventor of OPA (patented by UCL and licensed to Mallinckrodt Pharma). BPV, RK, AB are employees of Cyberliver Ltd. GM is a co-founder of Hepyx Ltd. **Author Contributions:** digital therapeutic design: GM, BPV, RK, AB, SA, KM, RJ; study concept and design: GM, KM, RJ; obtained funding: GM, KM; acquisition of data: GM, AN, SM, MS, MS, SRS, JK; analysis and interpretation of data: GM, SL, AN, LM; drafting of the manuscript GM, SL, SA, RJ; critical revision of the manuscript for important intellectual content: all authors.

## Abstract

**Background and aims:** Maintenance of abstinence in alcohol-related liver disease (ARLD) is a major unmet therapeutic need. Digital therapeutics can deliver ongoing behavioural therapy, in real-time, for chronic conditions. The aim of this project was to develop and clinically test AlcoChange, a novel digital therapeutic for ARLD.

**Methods:** AlcoChange was developed using validated behaviour change techniques (BCTs) and a digital alcohol breathalyser. This was an open-label, single-centre study. Patients with ARLD, ongoing alcohol use (within 1 month) and possession of a suitable smartphone were eligible. Patients were recruited from inpatient and outpatient settings, and received AlcoChange therapy for 3-months. The primary outcome was reduction in alcohol use from baseline to 3-months, measured by timeline follow-back (TLFB). Secondary outcomes included: (i) compliance with the AlcoChange app, (ii) alcohol-related and all-cause hospital re-admissions up to 1-year, (iii) qualitative analysis to determine factors associated with compliance.

**Results:** Sixty-five patients were recruited, of whom forty-one completed the study per-protocol. Patients compliant with the intervention (>60 logins over 3-months) had a significant reduction in alcohol use from baseline compared to non-compliant patients [median (IQR): −100% (100% to −55.1%) *vs* −57.1% (−95.3% to +32.13%), p=0.029]. The proportion attaining abstinence at 3-months was higher in the compliant group (57.1% *vs* 22.2%, p=0.025). The compliant group had a significantly decreased risk of subsequent alcohol-related re-admission up to 12-months (p=0.008). Qualitative analysis demonstrated receiving in-app feedback and presence of health-related ‘sentinel event’ were predictors of compliance with the intervention.

**Conclusions:** Use of the novel digital therapeutic, AlcoChange, was associated with a significant reduction in alcohol use and increase in proportion attaining abstinence in ARLD patients. Definitive, randomized trials are warranted for this intervention.

## Introduction

Globally, there is significant alcohol misuse and alcohol-related harm. There are 2.4 billion drinkers worldwide, and in 2016 alcohol use was associated with 3 million deaths (5.3% of all deaths), greater than hypertension and diabetes combined^1,2^. The major healthcare burden from alcohol misuse is the development of liver disease, which is now the second most common cause of preventable death in the UK^3^. Around 50% of liver-related deaths worldwide are due to excess alcohol, which increases to around 80% in the UK^1,4^.

Once liver disease is present, on-going alcohol use leads to markedly increased morbidity and mortality. Continued drinking following diagnosis leads to ∼50% mortality at 3 years, whereas with abstinence >75% are alive at seven years^5^. Therefore, maintaining abstinence is a key therapeutic goal in alcohol-related liver disease (ARLD). Alcohol rehabilitation therapy for patients with ARLD is associated with reductions in rates of hepatic decompensation and mortality^6^, yet less than one in five hazardous drinkers receive a behavioural intervention of any kind^7^.

Moreover, the Covid-19 pandemic has greatly exacerbated the healthcare impacts of alcohol-related harm. Emerging international data supports a rise in alcohol use – data from the US demonstrates an increase in overall alcohol consumption as well as heavy drinking days^8^. The impact of the pandemic on alcohol-specific morbidity and mortality is starker. Alcohol-related deaths increased by ∼25% in the USA between 2019 and 2020^9^, and a similar pattern was seen in the UK^10^. Moreover, the proportion of patients with ARLD being admitted with the most severe stage of the disease, termed acute-on-chronic liver failure, has also increased^11^.This has been attributed to the reduction in availability of outpatient clinics and alcohol services during the pandemic. As such, digital therapeutics are potentially a novel approach to remotely deliver monitoring and interventions to address this sharp upturn in morbidity and mortality.

The benefits of digital therapeutics in this context are on many levels, such as facilitating on-going clinical contact, allowing remote monitoring of the disease and providing scalable behavioural interventions in real time. However, there are few prospective studies exploring the feasibility and fidelity of a digital therapeutic for the management of chronic healthcare conditions in secondary care^12–14^, and none for the management of ARLD.

AlcoChange is a novel digital therapeutic, that has been developed specifically for patients with ARLD, delivered through a smartphone app and an associated breathalyser. AlcoChange was developed following feedback from focus groups and uses nudge theory and the health psychology theory of behaviour change to implement a series of behaviour change interventions (BCIs) to increase the likelihood of engaging in health-promoting and alcohol-reducing behaviours, such as monitoring of craving, alcohol consumption, measurement of breath alcohol and motivational messaging in response to patient triggers (Figure 1). Here, we report results of a proof-of-concept study of AlcoChange, in addition to standard of care, for patients with ARLD and ongoing alcohol use. The specific aims of this study were to assess feasibility of use, compliance with and efficacy of AlcoChange in eligible ARLD patients admitted to a specialist secondary and tertiary liver centre at the Royal Free Hospital, London and to qualitatively explore factors that influenced engagement with AlcoChange amongst ARLD patients.

**Figure 1.**
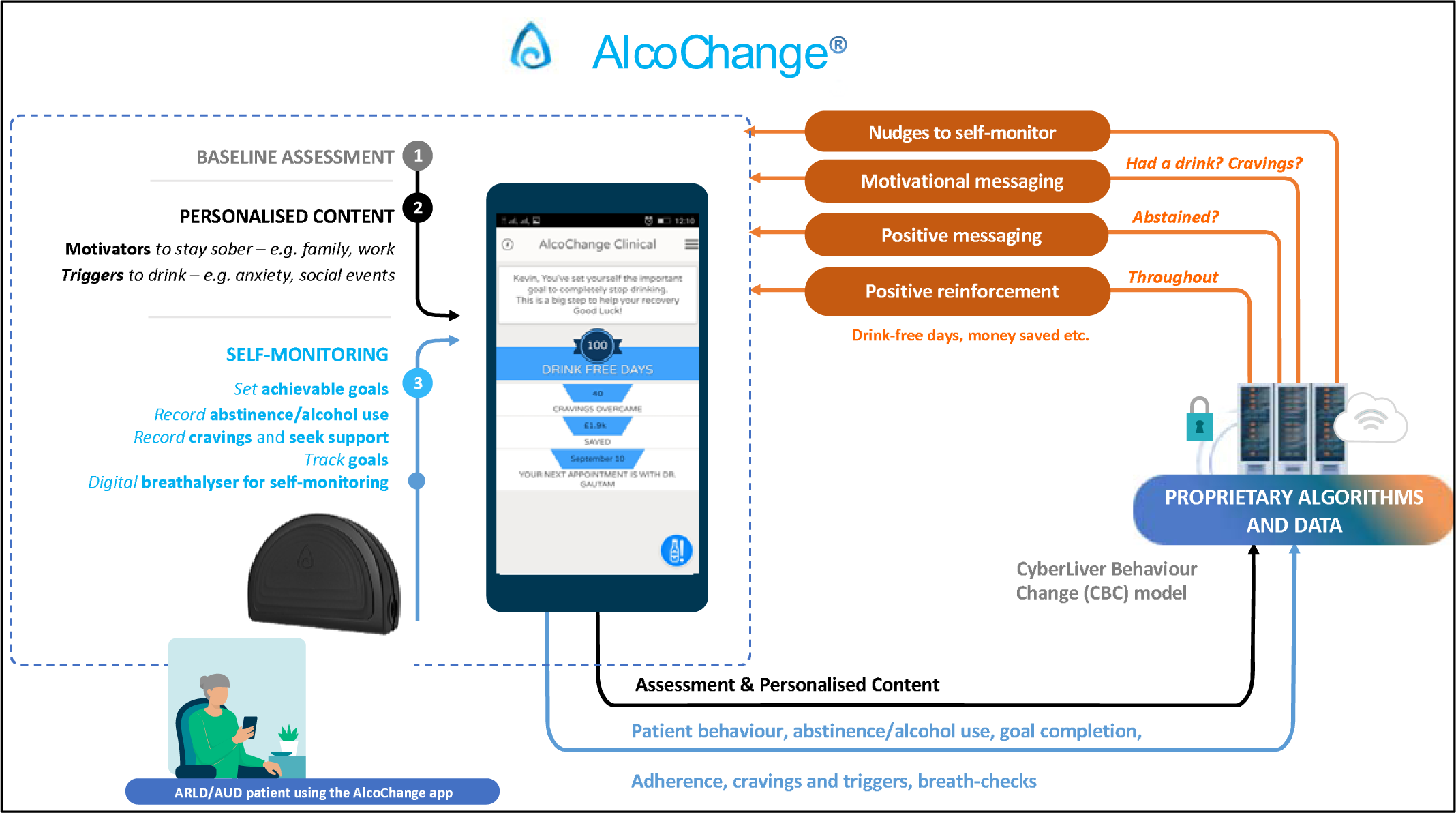
Outline of AlcoChange app and device functionality. The AlcoChange app utilises an automated behaviour change model. Personalised content is generated following a 30-60 minute baseline assessment, in response to real-time data (cravings, alcohol consumption etc.). This content is based on specific behaviour change techniques (BCTs) including: self-monitoring, positive reinforcement and goal-setting.

## Methods

### Study Participants

The study was conducted at Royal Free Hospital, London NHS Trust (ClinicalTrials.gov Identifier: NCT03474328), and was approved by the Research Ethics Committee (REC 16/SW/0035; protocol attached in Supplementary material). Patients with ARLD, defined by imaging features of chronic liver disease or clinical features of decompensated cirrhosis and a history of hazardous drinking, were eligible for inclusion. The other inclusion criteria were possession of a compatible smartphone, recent alcohol use (within 4 weeks) and reported intent to achieve abstinence. The initial exclusion criteria were Child-Pugh score >7 and inability to provide informed consent. The exclusion criterion of Child-Pugh score >7 was removed by ethics amendment, after 20 patients were recruited and evaluated for safety. All participants additionally received usual clinical care, which included review by a member of a multi-disciplinary ‘alcohol care team’ (ACT) and a brief inpatient behavioural intervention.

### Design of the Digital Therapeutic

Behaviour change techniques (BCTs) have been defined as a series of small, observable, replicable components with the potential to bring about behaviour change.^15^ Prior work from Abraham and Michie developed a taxonomy of BCTs for alcohol use disorder (AUD) and identified that ‘prompt self-recording’ of alcohol use was the most effective aspect for reduction of alcohol use.^16^ Further work from this group aimed to identify the optimal BCTs and strategies of engagement for inclusion in a smartphone app for AUD;^17^ twelve BCTs were identified as likely to be effective of which nine were used in the design of AlcoChange (Table 1, and Figure 2). Additionally, AlcoChange was developed with breath alcohol measurement hardware for self-monitoring and/or confirmation of abstinence. Data generated by the user was anonymised and stored in the NHS-compliant N3 server. These data were used to generate the digital behaviour change interventions (BCIs, see below) but were otherwise not shared with healthcare providers.

**Figure 2.**
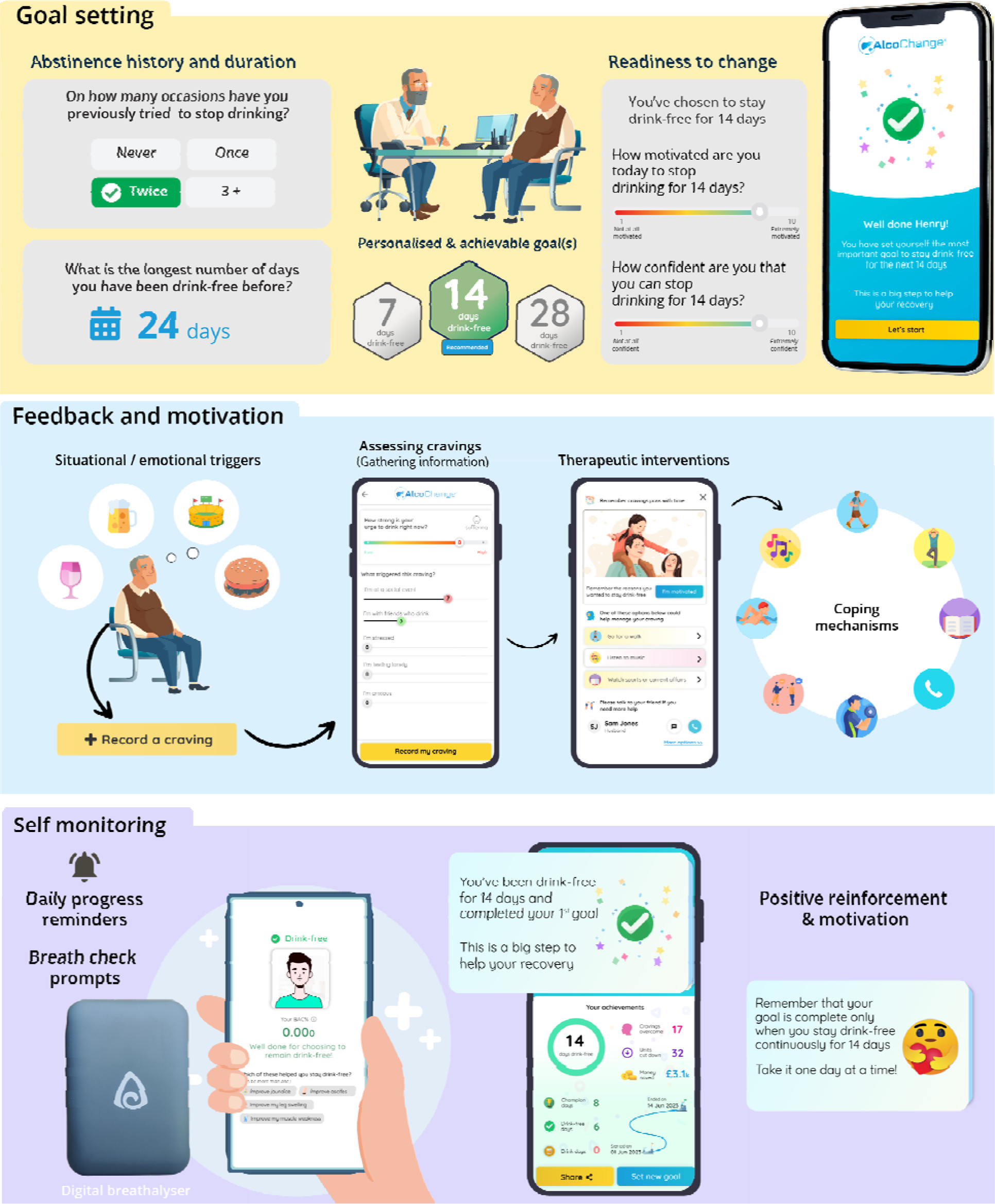
Examples of Behaviour Change Techniques (BCTs) delivered through AlcoChange. The AlcoChange app interacts with cloud-based platform to deliver behaviour change interventions (BCIs) in response to data collected in the app. Example BCTs and BCIs include ‘Goal setting’ *(top panel)*, ‘Feedback and motivation’ *(middle panel)* and Self-monitoring *(bottom panel)*.

**Table 1:** List of behaviour change techniques (BCTs) used in the AlcoChange app.

|  | **AlcoChange: Behaviour Change Techniques (BCTs)** |
| --- | --- |
| **BCT1** | ***Assessment and readiness to change*** |
| BCT1.1 | Assessment of drinking behaviour through AUDIT Questionnaire and alcohol history |
| BCT1.2 | Identify and document reasons for committing to abstinence |
| BCT1.3 | Assess level of motivation, confidence, readiness and ability to abstain or reduce alcohol consumption |
| BCT1.4 | Identify barriers to achieving abstinence (triggers for craving or alcohol consumption; situational, emotional and personal precipitants) |
| **BCT2** | ***Goal setting*** |
| BCT2.1 | Facilitate goal setting for abstinence |
| BCT2.2 | Prompt review of goals achieved and setting new, incremental targets |
| **BCT3** | ***Action planning*** |
| BCT3.1 | Help patient to identify and document the triggers that induce cravings or alcohol consumption |
| BCT3.2 | Direct the patient to coping strategies that may help overcome cravings, e.g. substitute activities, changes in routine |
| **BCT4** | ***Self monitoring and patient engagement*** |
| BCT4.1 | Prompt and facilitate self-recording of abstinence or alcohol use (recorded in-app or by breathalyser use) |
| BCT4.2 | Prompt and facilitate self-recording of commitment towards drink-free goals |
| BCT4.3 | Facilitate self-recording of cravings and environmental/situational triggers for cravings |
| BCT4.4 | Facilitate self-recording of successful craving management |
| **BCT5** | ***Provide information*** |
| BCT5.1 | Provide information on health consequences of alcohol use in liver disease |
| BCT5.2 | Provide information on benefits of abstinence in liver disease, and benefit of monitored abstinence over gradual reduction |
| BCT5.3 | Summarise commitments made and goals achieved |
| BCT5.4 | Provide information on benefits of self-monitoring and recording alcohol use and cravings |
| **BCT6** | ***Feedback and motivation*** |
| BCT6.1 | Boost confidence in the ability to abstain (boost motivation and self-efficacy) |
| BCT6.2 | Provide feedback on abstinence (progress report and calendar) |
| BCT6.3 | Provide positive feedback, praise, and demonstrate rewards (e.g. money saved, cravings overcome) |
| BCT6.4 | Affirm, or reaffirm, reasons to stay abstinent, and prompt patient |
|  | commitment |
| BCT6.5 | Provide positive feedback for progress towards abstinence goals |
| BCT6.6 | Provide reassurance when patients consume alcohol or fail to meet a goal |
| BCT6.8 | Prompt use of patient-selected motivational imagery |
| **BCT7** | ***Replacement activities*** |
| BCT7.1 | Suggest coping strategies and replacement activities at the time of craving recording |
| BCT7.2 | Provide advice on avoidance of triggers, and coping strategies to overcome triggers |
| **BCT8** | ***Social support*** |
| BCT8.1 | Provide options to seek additional support from friends and family |
| BCT8.2 | Provide access to alcohol helplines (e.g. UK Drinkline) |
| **BCT9** | ***Empathy/Normative feedback*** |
| BCT9.1 | Demonstrate empathy by responding to patient's perspective as understandable, comprehensible and valid |
| BCT9.2 | Accept ambivalence |
| BCT9.3 | Explain what is 'common' for most people in this situation |

The smartphone breathalyser was developed as a battery-less device to be paired with a smartphone (through a 3.5mm cable), rather than for independent use. This allows linkage of data with the AlcoChange app, and confirmation of identity of the user through activation of the front smartphone camera. The device was CE marked and approved by the Medicines and Health Regulatory Authority (UK) as an ‘*in vitro* diagnostic device’; quality assurance testing confirmed high correlation with blood alcohol measurements, and with breath alcohol measures from other commercially available breathalysers (Supplementary Document).

### Functionality of the AlcoChange app and platform

Patient data regarding health status, drinking patterns, triggers for craving and alcohol use, and motivational factors, are gathered during an extensive baseline assessment. These data are then processed by the cloud-based platform to deliver BCIs in real-time, in response to patient triggers. Specific examples of BCTs and interaction with the platform are listed below, and in Figure 2:

- **Readiness to change (BCT1)** and **Goal Setting (BCT2):** Assessing levels of motivation and confidence, and incremental goal setting throughout the treatment period, enhances self-efficacy and empowers patients to understand drivers for change and abstinence.
- **Self-monitoring (BCT4):** AlcoChange facilitates self-monitoring through a digital breathalyser to record blood alcohol content (BAC) and demonstrate abstinence, as well as monitoring of cravings and response to therapy.
- **Feedback and motivation (BCT6):** Personalised reminders are sent based on patient progress (goals achieved, cravings overcome etc). Empathy and normative feedback are used to boost motivation and facilitate engagement if goals are not met, and positive messaging is used when abstinence goals are achieved.
- **Replacement activities (BCT7):** In response to cravings, AlcoChange nudges the patient to assess and reflect on triggers for craving, and recommends personalised replacement activities and/or coping strategies based upon data gathered at baseline.

### Study Design

Recruitment for the study was conducted from both inpatients and outpatients at Royal Free London NHS Hospital Trust. Patients were recruited for the study from hepatology outpatients or following inpatient review by the alcohol care team prior to hospital discharge. After informed consent was obtained, baseline alcohol intake was assessed using the timeline follow-back method (TLFB), which is a psychometrically sound and widely-used tool for retrospective assessment of daily alcohol intake^18^. Patient empowerment with health services was quantified at baseline using the Support USers To Access Information and Services (SUSTAINS) questionnaire, and socio-economic status assessed using the English indices of deprivation tool.

Subsequently, participants received a 30-60 minute education session on download and use of the AlcoChange app and device. This included entering personalised ‘triggers’ for drinking and ‘motivators’ for abstinence, as outlined above. These motivators contributed to content for personalized motivational messages, which were delivered to the user in response to cravings or other triggers (Figures 1 and 2). Following this initial session, there was no specific intervention from the trial team until the follow-up visit at 3-months, although technical help for the app or breathalyser was available by email. Data from the app, such as alcohol use or compliance with the app/breathalyser, was only visible to the participant and not shared with the clinical team.

Since this was a proof of concept study, analyses were descriptive and exploratory. The primary outcome measure was reduction in self-reported alcohol intake at 3 months (10-14 weeks). This was assessed by a specific study visit at 3 months for assessment of recent alcohol use by TLFB. Participants were followed-up for 1-year following the 3-month treatment period. Additionally, a subgroup of participants was invited to attend facilitated focus groups for qualitative data collection after the follow up visits for the last patient recruited had been completed. Secondary outcomes included, (i) compliance with the app (anonymised and monitored remotely: number of logins, duration spent on the app); (ii) clinical outcomes assessed by alcohol-related and all-cause hospital re-admissions up to 1-year following the study period determined by comprehensive review of participant medical records and (iii) qualitative analysis to determine factors associated with compliance.

### Qualitative Assessment of Digital Therapeutic

Ethical approval for focus groups was obtained as a study amendment (amendment available in Supplementary material). Purposive sampling methods were used, targeting two respondent groups: compliant and non-compliant, defined by a threshold of logins from the previous stage of the study. Potential participants were contacted by telephone. Focus groups were conducted by AN and GM. Five compliant and two non-compliant participants took part in two focus groups. A semi-structured topic guide was developed based on a literature review, expert consultation, and preliminary findings from the pilot study; this was used to facilitate discussions (topic guide available in Supplementary material). Participants were asked motivations for participation in the study, expectations for the AlcoChange app/device, patterns of alcohol use and influences on past and current drinking behaviour, goals for alcohol use and abstinence, and experiences with AlcoChange including facilitators and barriers to use.

### Data Analysis

As an open-label, single group study, data was analysed per-protocol. Continuous variables were represented as mean ± standard deviation or median (interquartile range) and compared using the Student’s t-test. Categorical variables were expressed as counts (percentages) and compared using Chi-squared test or Fisher’s exact test. Area under the receiver-operating curve (AUROC) was constructed to determine the threshold for compliance/non-compliance in determining reduction in alcohol consumption. Hospitalisations were assessed at 1-year following the treatment period; these data were analysed using Kaplan-Meier analysis. All tests were two-tailed and a P value less than 0.05 was considered statistically significant. All analysis was conducted by R 3.6.2 (https://www.r-project.org/).

Qualitative data was evaluated using thematic analysis was used for qualitative data, following guidelines for best practice.^19^ Specifically, the following procedure was employed: 1) transcripts were read several times to increase familiarity with the data. Insightful comments made by the participants were noted; 2) initial codes were generated by pen and paper, i.e., labels were attached to important sections of discussion; 3) once the transcripts were coded, a provisional list of codes was created and sorted into potential overarching themes and subthemes; 4) the list of themes was reviewed and refined, the transcripts were revisited to explore whether each theme and subtheme satisfactorily represented the data; 5) a final list of theme and subthemes was defined and named.

## Results

### Patients

Consecutive patients admitted with ARLD to Royal Free London between April 2017 and March 2018, and referred to the study team by the alcohol care team were pre-screened for eligibility for the study. The flowchart of study recruitment is presented in Figure 3. Smartphone penetrance in this population was 64%. Sixty-five patients were recruited and consented for entry to the study. Forty-one (63%) participants completed the baseline and 3-month study visits. Eighteen patients (28%) were lost to follow-up, two (3%) had incomplete data sets, one was ineligible due to prolonged abstinence prior to admission (2%), one was unable to download the app due to smartphone issues (2%), and two (3%) died. Baseline characteristics of participants who completed both study visits are provided in Table 2, and characteristics of all enrolled patients are in Supplementary Table 1. Characteristics of patients completing the study were similar to the group who did not complete the study, aside from the proportion with cirrhosis which was significantly greater in the group who did not complete (Supplemental Table 1; 45.8% *vs* 22.0%, p=0.04). There was no difference in the proportion with decompensated cirrhosis between these groups.

**Figure 3.**
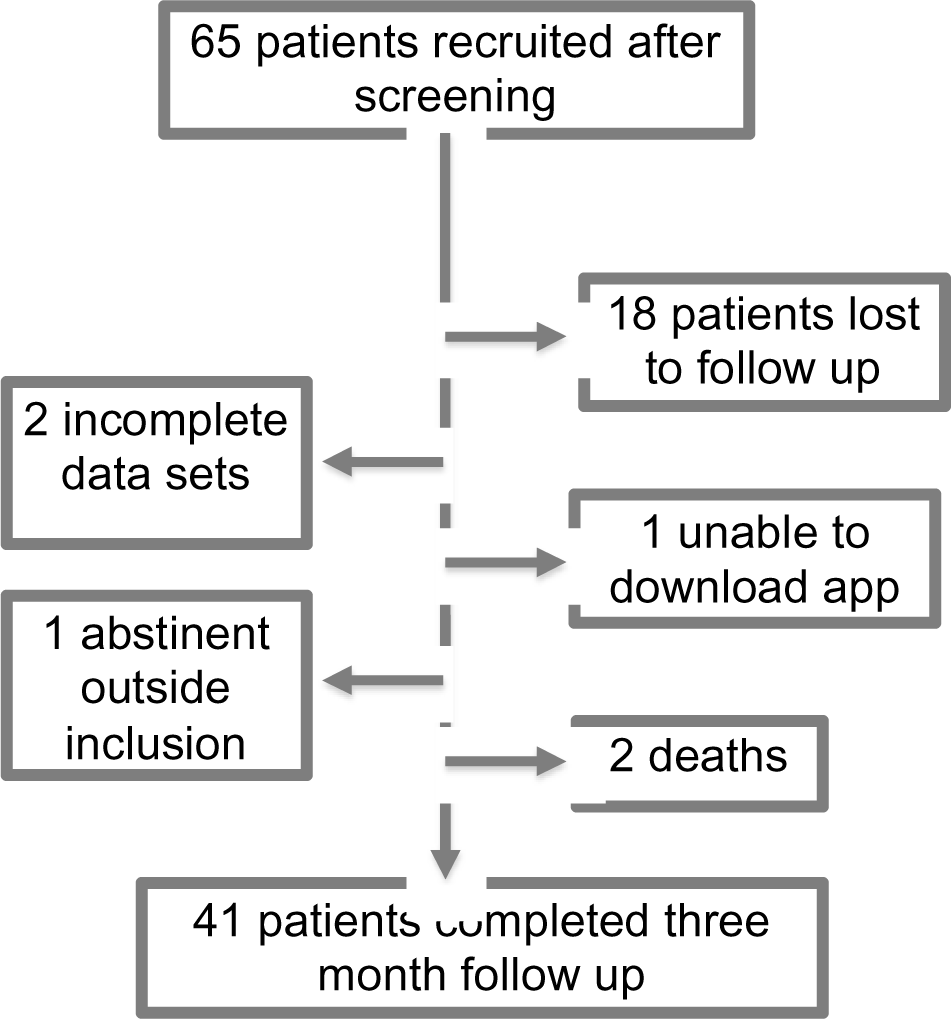
Flow-chart of patient recruitment.

**Figure 4:**
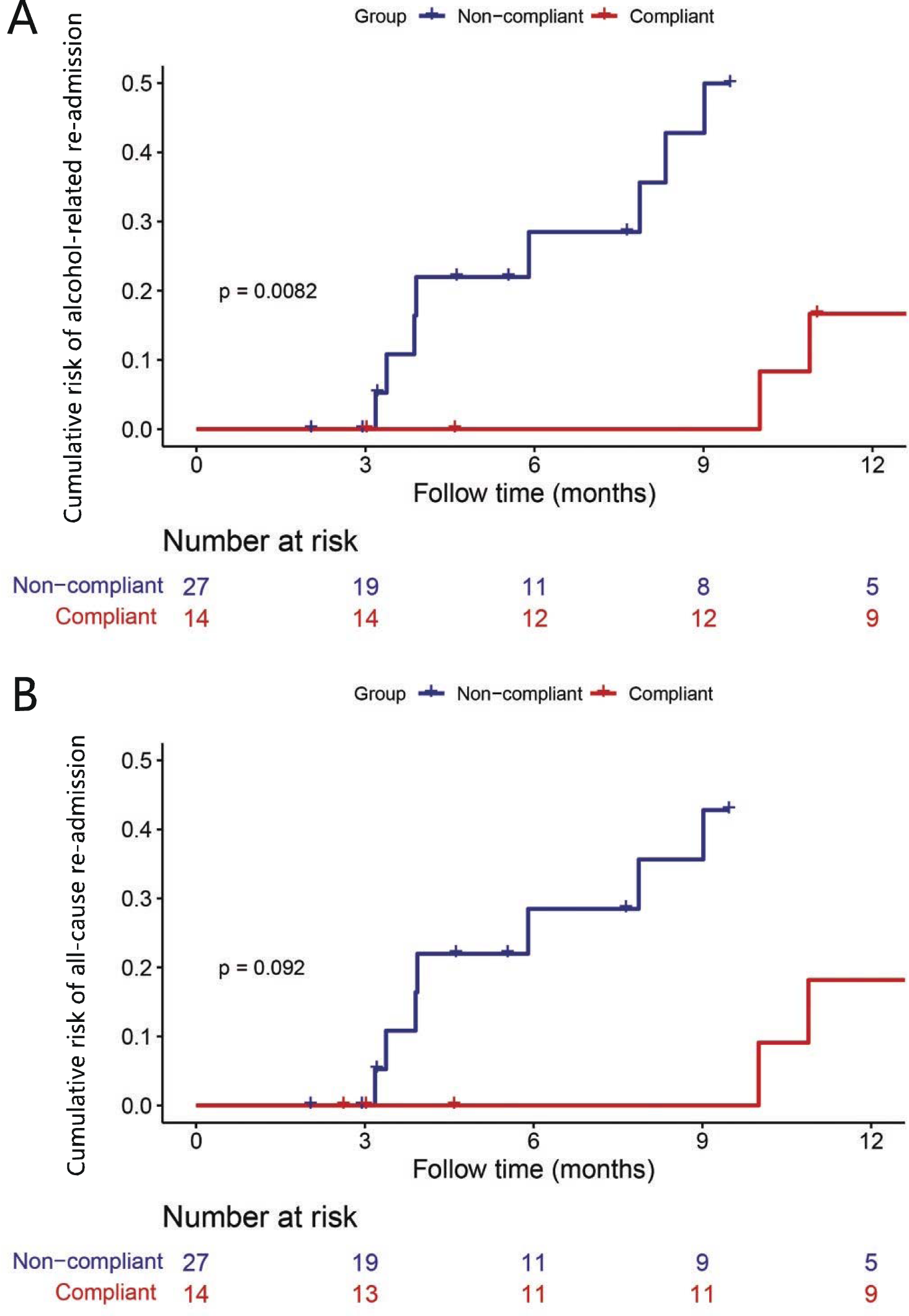
Cumulative risk of (A) alcohol-related re-admission, and (B) all-cause re-admission *(bottom panel)*, according to compliance/non-compliance. Risk of alcohol-related re-admission was significantly reduced in the compliant group compared with the non-compliant group in the 12-month follow-up period (*top panel*, Kaplan-Meier analysis, p=0.006).

**Table 2.**
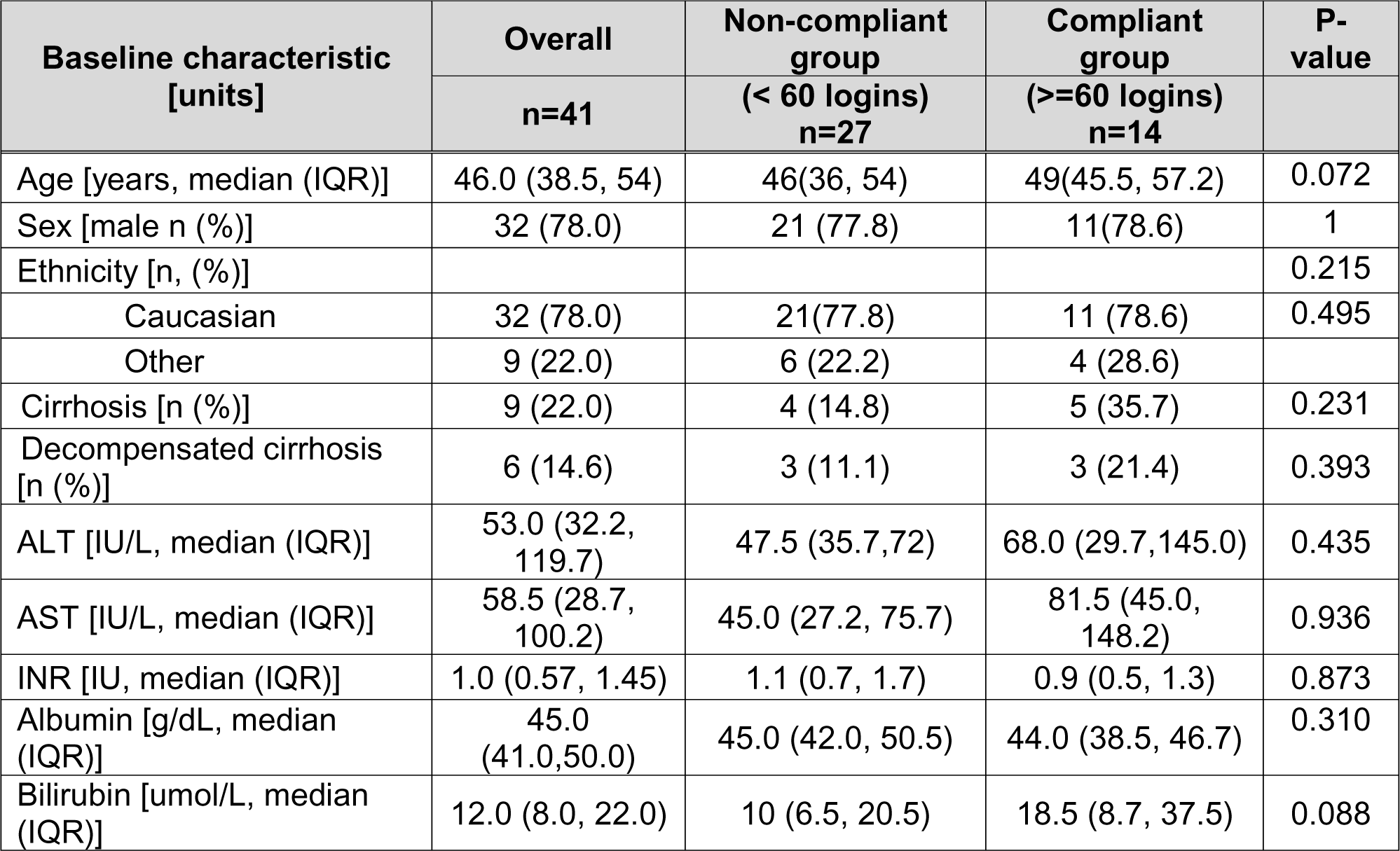
Demographic characteristics of participants completing baseline and 3-month visits.

### Self-reported alcohol intake

Self-reported alcohol intake for participants completing baseline and 3-month visits showed a significant reduction over the 3-month period, from 470.4g/week (254.4-896.0) to 84.8g/week (0-384.0) [median (IQR), p<0.01), table 3]. The AUROC of app login for determining abstinence at 3-months was 0.624 (95%CI: 0.452-0.797, p=0.176). The best cut-off value of app login determining abstinence at 3-months was 60 logins, with a sensitivity of 43.48% and a specificity of 77.78%. This approximates to use of the app four times each week. Applying this cut-off, a significant difference was noted in reduction in alcohol use between compliant and non-compliant groups [(median reduction (IQR) −100% (100% to −55.1%) *vs* −57.1% (−95.3% to +32.13%), p=0.029, Table 3]. Additionally, there was a significant difference between compliant and non-compliant groups in the proportion attaining complete abstinence at the end of therapy (57.1% *vs* 22.2%, p=0.025). There was a non-significant trend towards a dose-effect of app use towards abstinence at the end of therapy (Supplementary Table 2; 0-30 logins, 30-60 logins, >60 logins, p=0.057).

**Table 3:** Alcohol consumption data of participants completing baseline and 3-month visits.

| Alcohol intake | Overall | Non-compliant group | Compliant group | P-value |
| --- | --- | --- | --- | --- |
|  | n=41 | (< 60 logins) n=27 | (>=60 logins) n=14 |  |
| Baseline alcohol intake [g/week, median (IQR)] | 448.0 (156.9, 731.2) | 448.0 (160.0, 735.2) | 400.0 (0, 756.0) | 0.764 |
| 3-month alcohol intake [g/week, median (IQR)] | 678.4 (0, 384.0) | 117.6 (24.0, 66.4) | 0 (0, 205.6) | 0.244 |
| Change in alcohol intake at 3-months [% , median (IQR)] | -75.49 (-100.0, -8.3) | -57.1 (-95.3, 32,1) | -100.0 (-100.0, -55,1) | 0.029 |
| Abstinence at week 12 of therapy [n (%)] | 14 (34.1) | 6 (22.2) | 8 (57.1) | 0.025 |

Multivariable modelling was conducted to identify factors predicting abstinence in participants completing baseline and 3-month visits. No factors were found to attain statistical significance on logistic regression. (Supplementary table 3).

### Re-hospitalization

Rehospitalization rates for patients in the per-protocol analysis were collected up to 1-year following the 3-month treatment period. Endpoints assessed were all-cause hospital admission and alcohol-related hospital admission (Figure 3). Eleven patients (26.8%) needed 21 re-hospitalisations during the study period. All, except 4, of these readmissions were related to on-going alcohol abuse; the detail of each readmission is shown in Table 4. As shown in Figure 3, participants in the compliant group had a significantly lower risk of alcohol-related hospital admission compared with the non-compliant group (p=0.008), and a trend toward decreased risk of all-cause hospital admissions were observed (p=0.092). The number of patients needed to treat (NNT) to reduce one alcohol-related rehospitalization across the per-protocol cohort was calculated at 6.5 for the 3-month treatment period, and 12.2 for the 12-months following treatment.

**Table 4.**
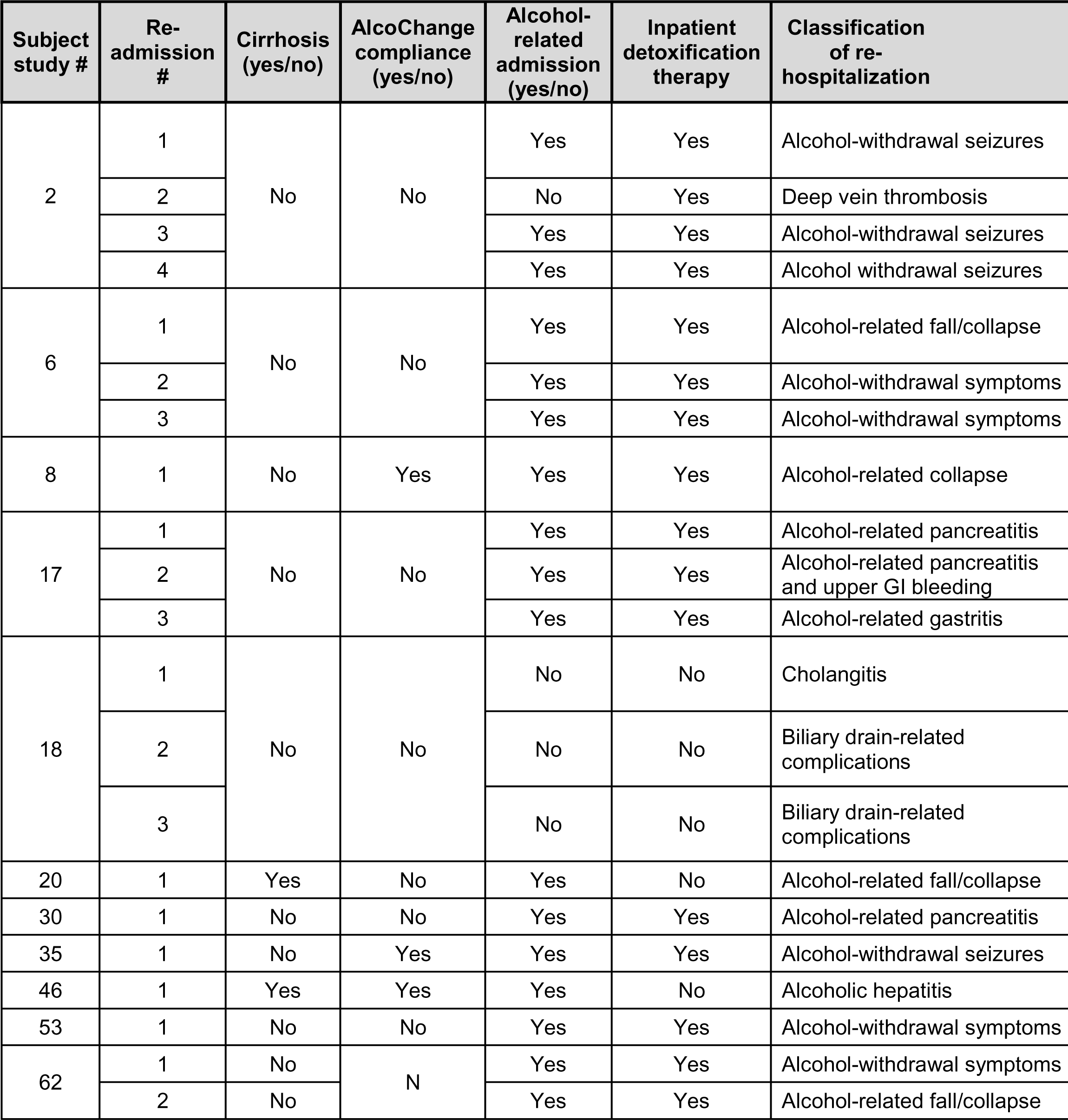
Detailed review of re-hospitalizations in patients completing the study. (Subject study numbers are linked-anonymised, and fully anonymised to individuals outside the research group)

| Subject study # | Re-admission # | Cirrhosis (yes/no) | AlcoChange compliance (yes/no) | Alcohol-related admission (yes/no) | Inpatient detoxification therapy | Classification of re-hospitalization |
| --- | --- | --- | --- | --- | --- | --- |
| 2 | 1 | No | No | Yes | Yes | Alcohol-withdrawal seizures |
|  | 2 |  |  | No | Yes | Deep vein thrombosis |
|  | 3 |  |  | Yes | Yes | Alcohol-withdrawal seizures |
|  | 4 |  |  | Yes | Yes | Alcohol withdrawal seizures |
| 6 | 1 | No | No | Yes | Yes | Alcohol-related fall/collapse |
|  | 2 |  |  | Yes | Yes | Alcohol-withdrawal symptoms |
|  | 3 |  |  | Yes | Yes | Alcohol-withdrawal symptoms |
| 8 | 1 | No | Yes | Yes | Yes | Alcohol-related collapse |
| 17 | 1 | No | No | Yes | Yes | Alcohol-related pancreatitis |
|  | 2 |  |  | Yes | Yes | Alcohol-related pancreatitis and upper GI bleeding |
|  | 3 |  |  | Yes | Yes | Alcohol-related gastritis |
| 18 | 1 | No | No | No | No | Cholangitis |
|  | 2 |  |  | No | No | Biliary drain-related complications |
|  | 3 |  |  | No | No | Biliary drain-related complications |
| 20 | 1 | Yes | No | Yes | No | Alcohol-related fall/collapse |
| 30 | 1 | No | No | Yes | Yes | Alcohol-related pancreatitis |
| 35 | 1 | No | Yes | Yes | Yes | Alcohol-withdrawal seizures |
| 46 | 1 | Yes | Yes | Yes | No | Alcoholic hepatitis |
| 53 | 1 | No | No | Yes | Yes | Alcohol-withdrawal symptoms |
| 62 | 1 | No | N | Yes | Yes | Alcohol-withdrawal symptoms |
|  | 2 | No |  | Yes | Yes | Alcohol-related fall/collapse |

The subgroup with underlying cirrhosis was small (n=9); no significant change in primary or secondary endpoints were found between compliant and non-compliant participants in this subgroup (Kaplan-Meier analysis, p=0.687).

### Qualitative study

The baseline outcomes from SUSTAINS questionnaire and socioeconomic status are presented in Supplemental table 4. There were no clear differences in patient empowerment between compliant and non-complaint groups. Indices of deprivation were also similar between compliant and non-compliant groups, aside from crime (rank and decile) which was lower in the non-complaint group.

Differences between compliant and non-compliant groups were further evaluated by qualitative assessment. Recruitment for the qualitative stage of the study was stratified on the basis of compliance or non-compliant groups but was otherwise random. The demographic characteristics of participants for this stage of the study are presented in Supplementary Table 3.

Across both compliant and non-compliant groups, several factors were raised which reportedly influenced engagement with the app and breathalyser. Specifically, experiences of feedback, sentinel events, and digital competency were felt to either facilitate or hinder app use; each theme is explored below.

#### Positive feedback

Receiving in-app feedback appeared to have acted as a psychological facilitator in promoting both continued abstinence as well as habitual use of AlcoChange. The money saving feature promoted the financial incentive of abstinence and was highlighted in particular by one app user who appreciated the ability to set goals. (quote A1, table 5). Motivational messaging resonated positively with another (quote A2, table 5) and finally, the emphasis on number of days sober built into the calendar strengthened engagement, particularly by sending daily reminders to use the breathalyser. This meant people continued to find a use for the app even during periods of sustained abstinence (quote A3, table 5)

**Table 5.** Focus group quotes regarding: (A) positive feedback from the app, (B) sentinel events and app compliance, and (C) digital competency.

| Quote reference | Representative quotes |
| --- | --- |
| **A1.** | *I'm the accountant so the one I liked best (feature of the app) was seeing the amount of money I'd saved stack up. That's my incentive. Well, I've saved three grand over the year that's good. I quite like that. I'm saving up for a car now (male aged 50-59, user)* |
| **A2** | *Not everyone is going to find it helpful you know it just sort of suits quirky people like me who a) aren't afraid of their phone and b) sort of quite like the 'yippee' thing (female aged 50-59, user)* |
| **A3** | *Well, I did find the reminder and I did use that everyday; so I now get a reminder. I do like the front page which says "well done you've completed these many days without a drink (male aged 40-49, user)* |
| **B1.** | *I mean I couldn't possibly dream of doing anything like giving her a fright like she had when I was in hospital (female aged 50-59, user)* |
| **B2** | *Because the shock of the seizure was so much, I didn't want to take any risks so I did everything that was available (male aged 50-59, user)* |
| **C1.** | *It [the app] just sort of suits quirky people like me who aren't afraid of their phone (male aged 50-59, user)* |
| **C2.** | *"The way the consultant explained to me you would be able to use your phone and I thought this was an interesting way of doing things, and I always use my phone" (female aged 40-49, user)* |
| **C3.** | *I did not understand it [the app] I kept trying to set everything up normal and I couldn't do it so I thought I probably should not bugger around with it in case I completely bugger it up (male aged 50-59, non-user)* |
| **C4.** | *It [the breathalyser] is not going to live for much longer I don't think. Sometimes you plug it in, and it says instantly 'blow now' and then it is not initialized so you have to take it out and start again (female, aged 50-59, user)* |
| **C5.** | *The cord is a bit short so when you put it in the top of the phone you have to squash in to kind of get it in your mouth, that could be improved (female, aged 40-49, user)* |

#### Sentinel event

A sentinel event is defined as an unanticipated event in a healthcare setting, resulting in death or serious physical or psychological injury to a patient. Three participants cited a sentinel event as being the catalyst for their desire for abstinence and subsequent engagement with the app. One participant described her event as a “*wake-up call*” and spoke of her desire to stay abstinent for her daughter (quote B1, table 5). Two participants were recruited whilst inpatients, recovering from variceal bleeding and an alcohol-related seizure, which appeared to encourage use of the app (quote B2, table 5).

#### Digital competency

Confidence in using a smartphone, as well as the average amount of time spent on a smartphone, appeared to have affected participants’ engagement with the app. Those who used the app regularly reported in the questionnaire that they would consider themselves to be “*very confident using a smartphone*” with all agreeing that they used their phone a “*medium to heavy*” amount. Both phone usage and confidence were specifically referenced when asked what about the study initially appealed to them (quote C1-2, table 5). Fear of technology, on the other hand, was raised by the two non-users, both of whom described themselves as “*not confident*” with their smartphone in the questionnaire. Additionally, feedback on the design and durability of the hardware was provided (quotes C3-6, table 5).

## Discussion

Smartphone-delivered digital therapeutics are a potentially effective and scalable way to remotely provide behavioural interventions for patients with ARLD. This proof-of-concept study made the novel observation that Alcochange, a digital therapeutic consisting of a smartphone app and digital breathalyser can be used successfully for self-monitoring of alcohol use and delivery of BCIs in patients with ARLD. Importantly, compliant patients also had a significant reduction in alcohol-related hospitalizations up to one year following the study period, although the intervention was only provided for up to 3-months, possibly indicating that the app can induce behaviour change. Moreover, there was a significant increment in the proportion of patients achieving complete abstinence from alcohol at the end of therapy in compliant patients, and a trend towards a ‘dose-effect’ of use of the app and abstinence.

This cohort is representative of the broader patient population with ARLD, being of working age (mean age 46.5), with male predominance, advanced liver disease in ≈20%, and high short-term mortality of over 13%/year. Using data for England in 2017-18 (NHS Digital), there were 1.2 million alcohol-related hospital admissions, the main age group for alcohol-related hospital admissions was 45–54 years, and 61% of admissions were male. Currently, smartphone penetrance in the UK is 85%, and the prevalence of smartphone use is likely to increase year-on-year. Around 35% of our cohort was compliant, and consequently this smartphone application has the potential to access this proportion of the high-risk ARLD population, with adequate engagement. This is in contrast to the face-to-face therapies that are intermittently received by less than 20% of this group^7,20^.

The key observations of this study are the significant reduction in alcohol consumption, and significant increase in the proportion of patients attaining complete abstinence, within the group compliant with app use. Importantly, this translated to a significant reduction in alcohol-related rehospitalization, with NNT=6.5 whilst on-treatment, and NNT=12.2 for the 1-year period following treatment. If these observations can be reproduced in real world practice, implementation of Alcochange would have considerable impact on healthcare costs.

Moreover, we proceeded to explore factors driving compliance and engagement with the app in our qualitative assessment. The focus groups revealed facilitators and barriers to app use. The ‘real-time’ nature of motivational messaging and positive feedback, particularly when out of hospital or during periods of abstinence, was considered an important factor promoting abstinence. Further, the focus groups suggested predictors of compliance and non-compliance. A sentinel event was considered an important positive predictor, suggesting that a smartphone intervention may be best delivered in an inpatient setting following a clinical event or soon after hospital discharge, rather than in the outpatient setting. As such, the intervention may be integrated into hospital ACTs rather than community services. By contrast, a lack of digital competency was a potential indicator of non-response. Additionally, the focus groups also revealed that hardware issues can be frustrating for this patient group, and any interventions requiring participants to use hardware must be robust to withstand varying lifestyles. Importantly these data have informed the development of the next generation AlcoChange 2.0, with features to enhance engagement in the ARLD population such as a Bluetooth-enabled breathalyser, and a physician-accessible dashboard to monitor compliance and abstinence. AlcoChange 2.0 is currently the subject of a randomized clinical trial (https://www.isrctn.com/ISRCTN10911773).

Patients with ARLD remain at high-risk of decompensation and death with on-going alcohol use^21^. Screening for alcohol use disorder amongst high-risk outpatients and inpatients, and referral to alcohol treatment professionals for this with established ARLD, is the clinical standard according to US guidelines^22^. A similar pathway exists in the UK for patients admitted with a complication of ARLD. However, UK data demonstrates that only 6% of individuals with harmful drinking received a behavioural intervention in the preceding 12 months^20^. Further, significant variations in care for patients with ARLD have been documented in the UK, with missed opportunities for intervention reported in patients with premature death due to ARLD^23^. Therefore, providing high-intensity alcohol support for individuals with ARLD is an unmet clinical need.

A recent retrospective study from the US suggests potential improvements in mortality with residential alcohol rehabilitation in ARLD^6^. However, the burden of ARLD has increased greatly following the Covid-19 pandemic, with an overall increase in mortality of ∼25%^9,10^. Consequently, residential and face-to-face interventions remain difficult to scale in the face of this rising tide of disease. The data presented here support digital therapeutics as a potentially useful and scalable modality to deliver behaviour change interventions in ARLD.

The limitations of this study are the lack of a control arm, which is justified as this is a first-in-man study of this digital therapeutic and the main aim of the study, which was feasibility and generating proof of concept data. All the analyses were, therefore, descriptive and the statistical analyses were exploratory, and the efficacy end points will need to be confirmed in future randomised clinical trials. Also, the single-centre nature of the study prevents claims of generalizability of the data and will need to be confirmed in future trials. Finally, although the dropout rate of 26% seems excessive, the population enrolled in this study have difficulties in compliance even with routine drug therapy and in clinical trials^24^. Nevertheless, these data demonstrate a dose-response relationship between the use of the device and abstinence, which was associated with clinically-relevant reduction in alcohol-related hospitalizations.

The Covid-19 public health emergency has highlighted the need for scalable solutions that allow remote monitoring and consultation for patients with chronic illness. Telemedicine is increasingly being utilized in many areas, including Hepatology, and carries many potential benefits such as delivery of targeted interventions in real time for vulnerable patients. However, prospective data to validate and direct the use of digital therapeutics in chronic disease management have been lacking. The proof-of-concept data presented here demonstrates that AlcoChange, digital therapeutic can be feasibly used for self-monitoring and delivery of BCIs in patients with ARLD in the acute setting and generate clinically-meaningful outcomes. Adequately powered, randomized, multicentre clinical trials are justified by these data, and have been initiated.

## Data Availability

All data produced in the present study are available upon reasonable request to the authors

## Notes

**Financial support.** This study was supported by a grant from The Health Foundation (Grant ref: 1943)

### Clinical Trial

ClinicalTrials.gov Identifier: NCT03474328

### Funding Statement

This study was supported by a grant from The Health Foundation (Grant ref: 1943)

### Author Declarations

This study was approved by the UK Health Research Authority (HRA) Research Ethics Committee (REC 16/SW/0035); protocol attached in Supplementary material).

