## Supplemental files for "Development and Clinical Evaluation of AlcoChange: a Digital Therapeutic for Patients with Alcohol-Related Liver Disease"

1. **Supplementary tables 1-5**
    - **Supplementary Table 1:** Characteristics of entire study cohort
    - **Supplementary Table 2:** Change in alcohol consumption in participants completing the study grouped by frequency of app use
    - **Supplementary Table 3:** Logistic regression for factors associated with abstinence in participants completing the study
    - **Supplementary Table 4:** Demographic characteristics of participants sampled for qualitative evaluation
    - **Supplementary Table 5:** Questionnaire data: *Education, Socio-economic status and Support USers To Access Information and Services (SUSTAINS)* in participants completing the study
2. **Study protocol and ethical amendments**
3. **Focus group topic guide**
4. **AlcoChange breathalyser calibration**

#### Supplementary Table 1.
##### Characteristics of entire study cohort (all patients recruited for participation)

| Baseline characteristic [units] | Overall | Completed study | Did not complete study | P-value |
|---|---|---|---|---|
| | n=65 | n=41 | n=24 | |
| Age [years, mean (SE)] | 46.1 (1.33) | 46.4 (1.7) | 45.6(2.0) | 0.773 |
| Sex [male n (%)] | 52 (80.0) | 32 (78.0) | 20 (83.3) | 0.607 |
| Ethnicity [n, (%)] | | | | |
| White | 52 (80) | 32 (78.0) | 20 (83.3) | 0.450 |
| Asian | 2 (4.9) | 2 (4.9) | 1 (4.2) | |
| Black | 6 (9.2) | 3 (7.3) | 3 (12.5) | |
| Other | 4 (6.2) | 4 (9.8) | 0 (0) | |
| Cirrhosis [n (%)] | 20 (30.8) | 9 (22.0) | 11(45.8) | 0.044 |
| Decompensated cirrhosis [n (%)] | 13 (20) | 6 (14.6) | 7 (29.3) | 0.157 |
| Ascites [n (%)] | 8(12.3) | 3(7.3) | 5(20.8) | 0.134 |
| HE [n (%)] | 0 | 0 | 0 | |
| ALT [IU/L, median (IQR)] | 49.0 (30.2, 105.0) | 53.0 (32.2, 119.7) | 48.5 (27.2, 84.0) | 0.435 |
| AST [IU/L, median (IQR)] | 59.0 (30.2, 93.7) | 58.5 (28.7, 100.2) | 64.0 (37.2, 87.0) | 0.936 |
| ALT/AST | 1.0 (0.6, 1.3) | 1.0 (0.6, 1.4) | 0.8 (0.6, 1.2) | 0.183 |
| INR [IU, median (IQR)] | 1.1 (1.0, 1.3) | 1.05(1, 1.1) | 1.1 (1.0, 1.3) | 0.266 |
| Albumin [g/dL, median (IQR)] | 45.0 (39.7, 47.2) | 45.0 (41.0, 50.0) | 33.0 (31.0, 47.0) | 0.036 |
| Bilirubin [umol/L, median (IQR)] | 13.0 (7.75, 22.2) | 12.0 (8.0, 22.0) | 14.0 (7.0, 26.0) | 0.080 |
| MELD score* [median (IQR)] | 8 (6, 10) | 8 (7, 10) | 7 (6, 10) | 0.731 |
| MELD-Na score* [median (IQR)] | 10 (7, 15) | 11 (7, 15) | 9 (6, 17) | 0.940 |
| CTP score* [median (IQR)] | 5 (5,5) | 5 (5,5) | 5 (5,7) | 0.090 |

***Abbreviations:*** Standard error (SE), hepatic encephalopathy (HE), alanine aminotransferase (ALT), aspartate aminotransferase (AST), international normalized ration (INR), model for end-stage liver disease (MELD), CLIF acute decompensation score (CLIF-AD), Child-Turcotte-Pugh (CTP).
*Disease scores only calculated for cirrhotic patients.

**Supplementary Table 2.**
**Change in alcohol consumption across study period grouped by frequency of app use**

| | | Number of login | | p-value |
|---|---|---|---|---|
| | (< 30 logins) n=18 | (30- 60 logins) n=9 | (>60 logins) n=14 | |
| Baseline alcohol intake [g/week, median (IQR)] | 303.2 (145.6-644.0) | 560.0 (433.6-1736.0) | 400.0 (0-756.0) | 0.098 |
| 3-month alcohol intake [g/week, median (IQR)] | 118.4 (28.8-384.0) | 84.8 (0-771.2) | 0 (0-205.6) | 0.230 |
| Change in alcohol intake at 3-months [%, median (IQR)] | -49.1 (-94.9, 43.5) | -64.9(-100, 13.1) | -100(-100, -55,1) | 0.124 |
| Abstinence at week 12 n (%) | 3(16.7) | 3(33.3) | 8(57.1) | 0.057 |
| Number of alcohol re-admission after study n (%): | | | | 0.188 |
| 0 | 13(72.2) | 5(55.6) | 12(85.7) | |
| 1 | 2(11.1) | 2(22.2) | 2(14.3) | |
| 2 | 0 | 1(11.1) | 0 | |
| >=3 | 3(16.7) | 1(11.1) | 0 | |

#### Supplementary Table 3.
##### Logistic regression for abstinence in participants completing the study

| | | Univariable | | | | Multivariable | |
|---|---|---|---|---|---|---|---|
| | OR | 95%CI | P | | OR | 95%CI | P |
| **Sex(male)** | 0.286 | 0.051-1.592 | 0.153 | | 0.210 | 0.020-2.173 | 0.191 |
| **Age** | 0.983 | 0.929-1.040 | 0.555 | | | | |
| **Cirrhosis** | 1.765 | 0.375-8.315 | 0.473 | | | | |
| **MELD score** | 1.198 | 0.934-1.537 | 0.155 | | 1.164 | 0.897-1.510 | 0.253 |
| **CTP score** | 1.622 | 0.641-4.104 | 0.307 | | | | |
| **Logins >60** | 2.692 | 0.675-10.740 | 0.161 | | 1.812 | 0.322-10.2 | 0.500 |

Variables with p< 0.2 in univariable analysis were selected for inclusion in multivariable analysis.
Method: backward, condition.

#### Supplementary Table 4:
##### Demographic characteristics of participants sampled for qualitative evaluation

| Baseline characteristic [units] | Range | n (%) |
|---|---|---|
| Age range [year] | 35 to 44 | 1 (14.3) |
| | 45 to 54 | 3 (42.9) |
| | 55 to 64 | 2 (28.6) |
| | 65 to74 | 1 (14.3) |
| Sex [male n (%)] | - | 5 (71.4) |
| Average weekly alcohol [units] | 0 | 5 (71.4) |
| | 1 to 7 | 0 |
| | 8 to 14 | 1 (14.3) |
| | 15 to 21 | 0 |
| | 22 to 28 | 1 (14.3) |
| Occupational status | Employed | 4 (57.1) |
| | Unemployed | 2 (28.6) |
| | Retired | 1 (14.3) |
| Education level | Not completed higher education | 2 (28.6) |
| | Completed higher education | 5 (71.4) |

**Supplementary Table 5.**
**Education, Socio-economic status and Support USers To Access Information and Services (SUSTAINS) questionnaire data**

| Questionnaire data | Overall | Non-compliant group | Compliant group | p-value |
|---|---|---|---|---|
| | n=41 | n=29 | n=14 | |
| **Education** | | | | 0.873 |
| Less than university | 23 (56.1) | 17 (58.62) | 6 (50) | |
| University or higher | 18 (43.9) | 12 (41.38) | 6 (50) | |
| **GP visit** | | | | 1 |
| >= once per month | 23 (56.1) | 16 (55.17) | 7 (58.33) | |
| < once per month | 18 (43.9) | 13 (44.83) | 5 (41.67) | |
| **Deprivation rank** | 16989.79 (1181.12) | 17312.48 (1513.99) | 16263.75 (1844.80) | 0.664 |
| **Deprivation decile** | 5.62 (0.36) | 5.78 (0.46) | 5.25 (0.57) | 0.509 |
| **Income rank** | 15169.77 (1147.40) | 15600.63 (1408.52) | 14200.33 (2026.49) | 0.576 |
| **Income decile** | 5.15 (0.35) | 5.26 (0.42) | 4.92 (0.62) | 0.653 |
| **Income score** | 0.12 (0.01) | 0.12 (0.01) | 0.13 (0.02) | 0.688 |
| **Employment rank** | 18201.64 (1338.36) | 18371.74 (1700.49) | 17818.92 (2170.90) | 0.843 |
| **Employment decile** | 6 (4,9) | 6 (4,9) | 5.5 (4,8) | 0.747 |
| **Employment score** | 0.08 (0.04, 0.1) | 0.08 (0.04, 0.1) | 0.08 (0.05, 0.1) | 0.843 |
| **Education rank** | 27355 (20311, 30218) | 27355 (19173.5, 30218) | 27291 (23777.25, 29521) | 0.879 |
| **Education decile** | 9(6,10) | 9(6,10) | 9(8,9) | 0.790 |
| **Health rank** | 23127.90 (1108.53) | 22933.81 (1403.60) | 23564.58 (1814.98) | 0.786 |
| **Health decile** | 8(6,9) | 8(6,9) | 8(6,10) | 0.734 |
| **Crime rank** | 12759.00 (1082.64) | 13633.67 (1487.36) | 10791.00 (941.25) | 0.115 |
| **Crime decile** | 4(3,6) | 4(3,7) | 4(3,4) | 0.247 |
| **Housing rank** | 10670.69 (917.85) | 10917.70 (1183.19) | 10114.92 (1404.53) | 0.666 |
| **Housing decile** | 3.67 (0.28) | 3.74 (0.37) | 3.50 (0.40) | 0.661 |
| **Environment rank** | 7684 (5547, 13088) | 8336 (6065.5, 11971.5) | 5952.5 (4868.25, 13332.5) | 0.394 |
| **Environment decile** | 3(2,4) | 3(2,4) | 2(2,4.75) | 0.446 |
| **How long does it take you to reach to the closest primary care centre? n (%)** | | | | 0.337 |
| No answer | 4 (9.76) | 4 (13.79) | 0 (0) | |
| 15 minutes | 22 (53.66) | 15 (51.72) | 7 (58.33) | |
| 15-30 minutes | 12 (29.27) | 9 (31.03) | 3 (25) | |

| | | | | |
|---|---|---|---|---|
| 30-60 minutes | 3 (7.32) | 1 (3.45) | 2 (16.67) | |
| **How long does it take you to reach to your general practitioner? n (%)** | | | | |
| No answer | 4 (9.76) | 4 (13.79) | 0 (0) | 0.573 |
| 15 minutes | 22 (53.66) | 15 (51.72) | 7 (58.33) | |
| 15-30 minutes | 12 (29.27) | 8 (27.59) | 4 (33.33) | |
| 30-60 minutes | 1 (2.44) | 0 (0) | 1 (8.33) | |
| 1-2 hours | 1 (2.44) | 1 (3.45) | 0 (0) | |
| More than 2 hours | 0 | 0 | 0 | |
| Don’t know | 1 (2.44) | 1 (3.45) | 0 (0) | |
| **How long does it take you to reach to closest hospital? n (%)** | | | | 0.387 |
| No answer | 4 (9.76) | 4 (13.79) | 0 (0) | |
| 15 minutes | 10 (24.39) | 5 (17.24) | 5 (41.67) | |
| 15-30 minutes | 14 (34.15) | 11 (37.93) | 3 (25) | |
| 30-60 minutes | 12 (29.27) | 8 (27.59) | 4 (33.33) | |
| 1-2 hours | 1 (2.44) | 1 (3.45) | 0 (0) | |
| More than 2 hours | | 0 | 0 | |
| Don’t know | | 0 | 0 | |

#### FULL/LONG TITLE OF THE TRIAL:

### AlcoChange: An Open Label Pilot Study of Smartphone Monitoring for Alcoholic Liver Disease

#### SHORT STUDY TITLE / ACRONYM

### AlcoChange

**This protocol has regard for the HRA guidance and order of content**

**RESEARCH REFERENCE NUMBERS:**

| | |
|---|---|
| **IRAS Number:** | 190682 |
| **SPONSOR / CO-SPONSORS / JOINT-SPONSORS:** | University College London |
| **SPONSORS Number:** | 15/0789 |
| **SITE:** | Royal Free London NHS Foundation Trust |
| **FUNDER:** | The Health foundation and CyberLiver |
| **FUNDERS Number** | The Health Foundation-CRM 1943 GIFTS 7470 |
| **PROTOCOL VERSION NUMBER AND DATE:** | Final v4.0 18th March 2018 |

#### SIGNATURE PAGE

The undersigned confirm that the following protocol has been agreed and accepted and that the Chief Investigator agrees to conduct the study in compliance with the approved protocol and will adhere to the principles outlined in Directive 98/79/EC of the European parliament and of the Council of 27th October 1998 on in vitro diagnostic medical devices and any subsequent amendments, device guidelines namely MEDDEV, the Sponsor's SOPs, and other regulatory requirements as amended.

I agree to ensure that the confidential information contained in this document will not be used for any other purpose other than the evaluation or conduct of the clinical investigation without the prior written consent of the Sponsor

I also confirm that I will make the findings of the study publically available through publication or other dissemination tools without any unnecessary delay and that an honest accurate and transparent account of the study will be given; and that any discrepancies from the study as planned in this protocol will be explained.

**For and on behalf of the Study Sponsor:**

Signature: ............................................................................................................

Date: ....../....../......

Name (please print): ............................................................................................................

Position: ............................................................................................................

**Chief Investigator:**

Date: 21/Jan/2016

Name:

Professor Kevin Moore

Position: Professor of Hepatology

**KEY TRIAL CONTACTS**

| | |
|---|---|
| Chief Investigator | Professor Kevin Moore<br>Professor of Hepatology<br>Royal Free London NHS Foundation Trust<br>Pond Street<br>London<br>NW3 2QG<br>Tel: 020 7794 0500 ext 36167<br> |
| Sponsor | University College London |
| Funder(s) | The Health Foundation<br>Will Warburton (Director of Improvement)<br>Registered Office: 90 Long Acre<br>London<br>WC2E 9RA<br><br>CyberLiver Ltd<br>Mr Bharad PV (Chief Technical Officer)<br>First Floor Burnells<br>5 Garland Road<br>Stanmore<br>Middlesex<br>HA71NR<br>United Kingdom |

| | |
|---|---|
| Key Protocol Contributors | Dr. Gautam Mehta<br>Locum Consultant/Senior Research and teaching fellow<br>University College London<br>UCL Institute of Liver and Digestive health<br>Royal Free Campus<br>London<br>NW3 2PF<br>Tel: 07715232734<br><br><br>Professor Kevin Moore<br>As above<br><br>Professor Rajiv Jalan<br>Professor of Hepatology<br>University College London<br>UCL Institute of Liver and Digestive health<br>Royal Free Campus<br>London<br>NW3 2PF<br>Tel: 02074332795<br> |
| Statistician | Yu-Mei Ruby Chang<br>Lecturer in Statistics<br>Research Support Office<br>The Royal Veterinary College<br>Royal College Street<br>London<br>NW1 0TU<br>Tel: 02032148008<br> |

**TRIAL SUMMARY**

| | | |
|---|---|---|
| Trial Title: | AlcoChange: An Open Label Pilot Study of Smartphone Monitoring for Alcoholic Liver Disease | |
| Internal ref. no. (or short title) | AlcoChange | |
| Trial Design | Open label, pilot | |
| Trial Participants | Inpatients and outpatients with alcoholic liver disease (ALD) | |
| Planned Sample Size | 60 | |
| Treatment duration | 3 months | |
| Follow up duration | Until end of treatment | |
| Planned Trial Period | 1 year | |
| | **Objectives** | **Outcome Measures** |
| Primary | Self-reported alcohol intake | Alcohol intake by the timeline follow-back method |
| Secondary | - Compliance with the device.<br>- Attendance at community alcohol services following discharge.<br>- Change in health related quality of life (HRQoL)<br>- Changes in patient empowerment<br>- Factors determining compliance with app and effect on drinking behaviour | - No of interactions with the device over the study period.<br>- Follow-up rates at community alcohol services appointments over the study period.<br>- HRQoL will be assessed by SF-36 questionnaire before and after the study period.<br>- Patient empowerment will be assessed by the SUSTAINS questionnaire.<br>- Qualitative assessment at focus group |
| Tertiary / exploratory | - Presence or absence of periodontitis in patients with ALD<br>- Change in microbial population in saliva and stool over study period<br>- Frequency of hospital admissions up to 1 year due to | - Dental examination at baseline<br>- Sampling of saliva and stool at baseline and at 3 months<br>- Hospital Episode Statistics (HES) data to determine subsequent hospital admissions up to 1 year. |

| | |
|---|---|
| | a complication of ALD. |

| Study Intervention: | AlcoChange is a smartphone application and battery-less smartphone breathalyser. AlcoChange (hereafter referred to as *the device)* is CE marked as a class I *in vitro* diagnostic medical device under Directive 98/79/EC (IVDD). |
|---|---|

**FUNDING AND SUPPORT IN KIND**

| **FUNDER(S)**<br>(Names and contact details of ALL organisations providing funding and/or support in kind for this trial) | The Health Foundation **(**£ 74,920.06) |
|---|---|
| | CyberLiver (manufacturing costs of the device) |

#### ROLE OF STUDY SPONSOR AND FUNDER

The sponsor for this study is UCL. The role of the sponsor is to ensure arrangements are provided for initiation, management and monitoring of the study. The sponsor will also ensure that an appropriate research ethics committee has reviewed the study and provided a favourable opinion, and that arrangements proposed for the work are consistent with this research governance framework.

The funder of this study is The Health Foundation and CyberLiver (manufacturer).

Neither the sponsor, nor the external funder, have any role in the trial design, data analysis, interpretation, manuscript preparation or dissemination.

#### ROLES AND RESPONSIBILITIES OF TRIAL MANAGEMENT COMMITTEES/GROUPS & INDIVIDUALS

Since this a small, single-centre pilot study, a Trial Management Committee will not be involved. However, patient involvement has been sought in the design of this study, with active engagement with Camden Alcohol Users Group to identify patient-reported outcomes that are assessed in the questionnaire data in the trial design. Day to day management of the study will remain the responsibility of the CI and the PIs.

The ethics application and submission and development of informed consent documents will be undertaken by an independent contract research organisation PHARMExcel Ltd.

Monitoring will also be performed by Pharmexcel Ltd, at a single time point during the study period.

##### Protocol contributors:

The protocol has been developed by Dr. Gautam Mehta, Professor Kevin Moore and Professor Rajiv Jalan. The Protocol has been reviewed by The Health Foundation as part of the funding review process, and UCL R&D department as part of the sponsorship process.

**KEY WORDS:**

Alcoholic liver disease

Alcohol

Smartphone

Digital health

Behaviour change

**LIST of CONTENTS**

| GENERAL INFORMATION | Page No. |
|---|---|
| TITLE PAGE | 1 |
| RESEARCH REFERENCE NUMBERS | 2 |
| SIGNATURE PAGE | 3 |
| KEY TRIAL CONTACTS | 4-5 |
| TRIAL SUMMARY | 6-7 |
| FUNDING | 7 |
| ROLE OF SPONSOR AND FUNDER | 8 |
| ROLES & RESPONSIBILITIES OF TRIAL MANAGEMENT COMMITTEES, GROUPS AND INDIVIDUALS | 8 |
| LIST of CONTENTS | 9 |
| LIST OF ABBREVIATIONS | 10-11 |
| TRIAL FLOW CHART | 12 |
| **SECTION** | |
| 1. BACKGROUND | 13 |
| 2. RATIONALE | 13-15 |
| 3. OBJECTIVES AND OUTCOME MEASURES/ENDPOINTS | 15-16 |
| 4. TRIAL DESIGN | 17 |
| 5. ELIGIBILITY CRITERIA | 17 |
| 6. TRIAL PROCEDURES | 18-22 |
| 7. DEVICE | 22-24 |
| 8. SAFETY REPORTING | 24-31 |
| 9. STATISTICS AND DATA ANALYSIS | 31 |
| 10. DATA HANDLING | 31-32 |
| 11. MONITORING, AUDIT & INSPECTION | 32 |
| 12. APPENDICIES | 33 |

**LIST OF ABBREVIATIONS**

| | |
|---|---|
| AE | Adverse Event |
| ALD | Alcohol Liver Disease |
| AR | Adverse Reaction |
| BCI | Behaviour Change Interventions |
| CA | Competent Authority |
| CI | Chief Investigator |
| CRF | Case Report Form |
| CRO | Contract Research Organisation |
| EC | European Commission |
| EMEA | European Medicines Agency |
| EU | European Union |
| GCP | Good Clinical Practice |
| HES | Hospital Episode Statistics |
| HRQoL | Health-Related Quality of Life Questionnaire |
| ICF | Informed Consent Form |
| ICH | International Conference on Harmonisation of technical requirements for registration of pharmaceuticals for human use. |
| ISF | Investigator Site File |
| ISRCTN | International Standard Randomised Controlled Trials Number |
| IVD | *In vitro* diagnostic |
| IVDD | *In vitro* diagnostic Directive |
| MHRA | Medicines and Healthcare products Regulatory Agency |
| MDD | Medical Device Directive |
| MS | Member State |
| NHS R&D | National Health Service Research & Development |
| PI | Principal Investigator |
| PIC | Participant Identification Centre |
| PIS | Participant Information Sheet |

| | |
|---|---|
| QA | Quality Assurance |
| QC | Quality Control |
| RCT | Randomised Control Trial |
| REC | Research Ethics Committee |
| RFH | Royal Free Hospital |
| SAE | Serious Adverse Event |
| SOC | Standard of Care |
| SAR | Serious Adverse Reaction |
| SDV | Source Data Verification |
| SOP | Standard Operating Procedure |
| SSI | Site Specific Information |
| SUSAR | Suspected Unexpected Serious Adverse Reaction |
| TLFB | Time Line Follow Back Method |
| TMG | Trial Management Group |
| TSC | Trial Steering Committee |
| TMF | Trial Master File |
| UCL | University College London |
| UNC | University of North Carolina |

#### TRIAL FLOW CHART

##### 1. Patient Flow

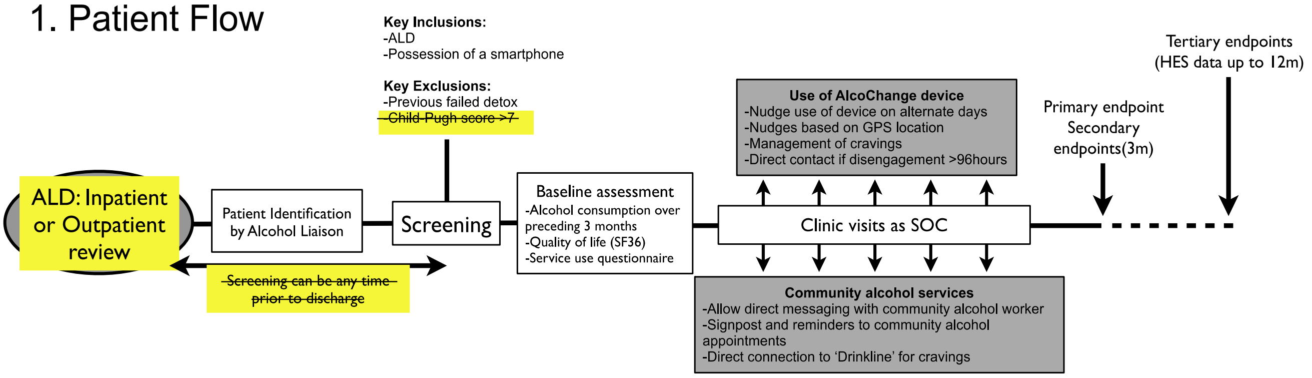

##### 2. AlcoChange messaging flowchart

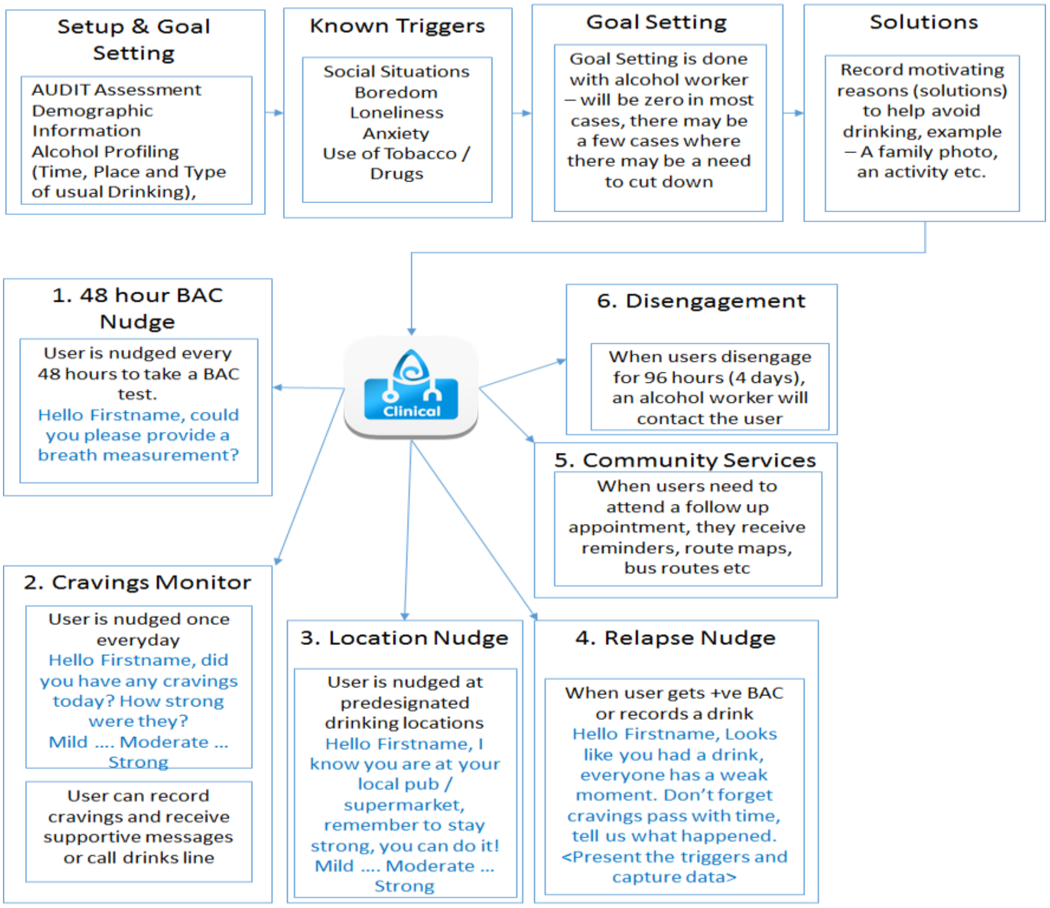

**STUDY PROTOCOL**

**AlcoChange: An Open Label Pilot Study of Smartphone Monitoring for Alcoholic Liver Disease**

#### 1 BACKGROUND

Alcohol misuse is a common cause of morbidity and mortality in the UK, accounting for more premature deaths amongst individuals of working age than cigarette smoking. It is estimated that 24% of the UK adult population drink at harmful levels, but only 6% of these receive any input from alcohol services. Abstinence leads to markedly improved outcomes in patients with alcoholic liver disease (ALD) - continued drinking following diagnosis leads to ~50% mortality at 3 years, whereas with abstinence >75% are alive at 7 years. Thus, **any intervention that decreases alcohol use and improves abstinence will improve mortality in this group**. Currently, there is no provision of high-intensity alcohol support for patients discharged with ALD, despite their high mortality.

**Behaviour change interventions (BCIs) are effective tools for reducing alcohol consumption**. Behaviour change interventions (BCIs) are effective at reducing alcohol consumption in targeted individuals, and have been the subject of over 20 randomised clinical trials (RCTs), and a Cochrane review(1). The most important aspect of a BCI that leads to decreased alcohol use is **self-monitoring of alcohol intake**. Thus, from systematic review, self-monitoring of alcohol is the single key technique that significantly predicts success of a brief intervention (1).

The aim of this pilot study is to provide 60 ALD patients with the AlcoChange device, to enable accurate self-monitoring of alcohol use, and determine changes in self-reported alcohol use over the study period.

1. Michie S et al. Identification of behaviour change techniques to reduce excessive alcohol consumption. Addiction 2012;107(8):1431-40.

#### 2 RATIONALE

Mobile phone technology has been previously demonstrated to change health behaviours - text messaging has been successfully used to reduce cigarette smoking(1). However, smartphones have not previously been used to facilitate BCIs in ALD, or to facilitate self-monitoring in any population with alcohol misuse.

The hypothesis of this study is that use of the AlcoChange device, according to the functionality outlined above in figure 2, will lead to a >20% reduction in self-reported alcohol intake, or a maintenance of abstinence, over a 3-month period in patients with ALD.

Since this is a pilot study, the study design is open-label, single-arm with no control group. Compliance with the device, as well as patient reported outcomes, quality of life and patient empowerment scores will be important secondary endpoints. A further endpoint will be qualitative assessment by focus group to explore: (i) facilitating factors and barriers to use of the app, and (ii) factors which influenced drinking behaviour in the study cohort.

Additionally, this study will incorporate tertiary, exploratory endpoints to address the role of oral health, and oral and gut microbes, in the natural history of ALD. A baseline dental examination will be performed, and baseline samples of saliva and stool will be collected to determine microbial populations. Saliva and stool samples will also be collected at the end of study visit, to determine changes in oral and gut microbe populations with decreased alcohol use. Data on hospital re-admission will be obtained from hospital episode statistics (HES) data to determine if baseline dentition or oral/gut bacterial populations predict re-admission in ALD.

1. Free C et al. Smoking cessation support delivered via mobile phone text messaging (txt2stop): a single-blind, randomised trial. Lancet 2011;378:49-55

#### 2.1 Assessment and management of risk

This study does not alter the standard of care (SOC) of patients admitted with ALD. The only intervention is a smartphone application and breathalyser (device), which has been CE marked as an *in vitro* diagnostic medical device, and hence the risks involved are minimal compared to SOC and are likely to minimise harm.

**Key Risks and Mitigation**

The main risks involved in the study and their respective mitigation are as follows,

1. **False positives breathalyser readings**
   In remote cases, it is possible that presence of alcohol is shown when in reality the user has not consumed any alcohol. The reasons for this could be ethanol residues from previous tests, ethanol generated by the human body, or consumption of cough syrups. In such cases, the risk will be managed through education and training. Users know that they have not consumed alcohol and will be informed that repeated tests can flush out accurate results.
2. **False negative breathalyser readings**
   In remote cases, it is possible that while the user has alcohol present in their blood, the reading may be displayed as nil. The reason for this may be due to inaccurate breath sampling (breath from the mouth as opposed to the lungs of the user). This risk will be mitigated through provision of appropriate risk messages (see below) even when the readings are nil.

**Warnings on Drinking and Driving**

Users will be strictly told not to drive after consuming any amount of alcohol. The message will be propagated in various sections of AlcoChange app where relevant as well as in a printed format along with the Breathalyser hardware.

**The following message will be shown in the "About Us" screen in the app:**

"Warning:
AlcoChange is not intended to be a medical device. You should consult a qualified medical professional for all matters concerning your health. AlcoChange cannot be held responsible for any actions by the user."

**The following message will be shown in the breathalyser results screen:**

"It is an offence to drive or perform any activity that may require you to be alert after consuming an alcoholic beverage, whatever the level of alcohol in the blood. Please note that the level of alcohol is not easily translated into the number of drinks you have had, since this depends on a variety of factors including gender, height, weight, stomach emptiness, type of drink and others. Any amount of alcohol will affect your ability to drive. Therefore it is best to never drink and drive. If you drive after a drink, you could be prosecuted and possibly convicted."

### 3 OBJECTIVES AND OUTCOME MEASURES/ENDPOINTS

#### 3.1 Primary objective / outcome measures

The primary objective is to determine the effect of use of the AlcoChange device on self-reported alcohol use at 3 months, amongst patients with ALD. The hypothesis is that use of the AlcoChange device will lead to a >20% reduction in self-reported alcohol use at 3 months.

The outcome measure of alcohol intake within the preceding 3 months will be assessed by the timeline follow-back method, which is a commonly used endpoint in alcohol intervention studies. This will be assessed by paired t-test or Wilcoxon signed-rank test.

The initial interview will be conducted in either the inpatient or outpatient setting, after the patient has been screened and consented for entry into the study. The second interview will be conducted in the outpatient setting, at the time of the patient's follow-up appointment at 3 months (between 10-14 weeks, as part of SOC).

#### 3.2 Secondary objectives / outcome measures

The secondary objectives will assess compliance with the AlcoChange device, attendance at community alcohol services, and assessment of health related quality of life (HRQoL) and patient empowerment.

Compliance with the AlcoChange device will be assessed by quantifying the number of interactions with the device by the patient over the 3-month study period.

Attendance at community alcohol services following discharge from hospital will be assessed by direct contact with the community alcohol centre[1] at the end of the study period (list of community

[1] Note the research team will contact the community alcohol services directly at the end of the study to gather information on community attendances. The community services will not have any involvement in the study or any access to study information.

alcohol centres attached). Current non-attendance rates following discharge from hospital are ~50%.

Health-related quality of life (HRQoL) and patient empowerment will be assessed by questionnaires, which will be administered at baseline and at the end of study follow-up appointment at 3 months. The SF-36 questionnaire will be used to assess HRQoL, and patient empowerment will be assessed by the SUSTAINS questionnaire which is designed and validated to assess the impact of e-health interventions on patient empowerment and relationship with health services.

Qualitative research methodologies will also be used in order to gain deeper understanding of facilitating factors and barriers to use of the app, and factors which influence drinking behaviour. Study participants will be contacted by telephone, and asked for their willingness to participate in a focus group to explore the above issues. Two focus groups will be conducted, with four to six participants in each group. Each will last up to one hour and a half. A topic guide will be developed to direct the focus groups. Participants will be asked for their thoughts about facilitators and barriers to use of the AlcoChange app, and factors that influenced changes in drinking behaviour. Specific interactions between use of the app and factors that influenced changes in drinking behaviour. Specific interactions between use of the app and changes in drinking behaviour will be explored.

##### 3.3 Exploratory endpoints/outcomes

There is increasing interest in the role of oral and gut microbes in the natural history of liver disease. Additionally, periodontal disease has been associated with adverse health outcomes in diabetes mellitus and coronary heart disease, through effects of endothelial function. Importantly, endothelial dysfunction is a key component of the development of complications of cirrhosis.

The hypothesis of this part of the study is that the presence of periodontitis, or the baseline oral and gut bacterial populations, will be associated with outcomes from liver disease with one year. No individual patient data will be acquired beyond the 3-month study period, but re-admission will be obtained from Hospital Episode Statistics. Additionally, the effect of reduced alcohol intake from baseline on oral and gut microbes will be assessed, by repeat saliva and stool sampling at 3-months.

The presence of periodontitis will be assessed by dental examination, which will be conducted during the patient’s inpatient stay. This will include a basic periodontal examination which should not exceed 15 min in duration. Using a manual University of North Carolina (UNC-15) periodontal probe, the tip of the probe is gently inserted into the gingival pocket and the depth of insertion measured in millimetres on the probe. Probing force should not exceed 20-25 gm. Further details of the dental examination are attached.

| Objectives | Outcome Measures | Timepoint(s) of evaluation of this outcome measure (if applicable) |
|---|---|---|
| **Primary Objective** | Self-reported alcohol intake will | Self-reported alcohol intake will |

| | | |
|---|---|---|
| - To compare self-reported alcohol intake amongst patients with ALD, following 3 months of use of the AlcoChange device. | be assessed at baseline and at 3 months by the timeline follow back method. Self-reported alcohol intake will be assessed for the preceding 3 months at both time points. | be assessed at baseline, as an inpatient, and again at the end of study visit at 3 months (10-14 weeks). |
| **Secondary Objectives**<br>- To assess compliance with the AlcoChange device.<br>- To assess attendance at community alcohol services following discharge.<br>- To assess changes in health related quality of life (HRQoL) during the study period.<br>- To assess changes in patient empowerment during the study period.<br>- Factors determining compliance with the app and effect on drinking behaviour | - Compliance will be assessed by analysing the number of interactions with the device over the study period.<br>- Follow-up rates at community alcohol services appointments over the study period will be assessed.<br>- HRQoL will be assessed by SF-36 questionnaire before and after the study period.<br><br>- Patient empowerment will be assessed by SUSTAINS questionnaire before and after the study period.<br><br>- Qualitative analysis by focus group of up to 12 participants | - Compliance will be recorded by the AlcoChange platform throughout the study period.<br><br>- Attendance at community alcohol follow-up will be assessed by direct contact with the community alcohol centre at the end of the study period.<br><br>- Questionnaires for HRQoL and patient empowerment will be assessed at baseline, as an inpatient, and again at the end of study visit at 3 months (10-14 weeks).<br><br>- Following completion of the 3 months period of use of AlcoChange |
| **Tertiary Objectives**<br>- To determine the prevalence of periodontitis amongst patients with ALD.<br>- To determine changes oral and gut microbial populations over the study period.<br>- To determine if the presence of periodontitis or saliva/stool microbial populations are related to frequency of hospital admissions up to 1 year. | - The presence of periodontitis will be determined by dental examination at baseline.<br>- Microbial populations in saliva and stool will be determined at baseline and at the end of the study period.<br>- The presence of periodontitis and microbial populations in saliva and stool will be correlated with frequency and type of readmission with liver disease up to 1 year. | - Dental examination will be conducted at baseline, as an inpatient.<br>- Saliva and stool sampling will be conducted at baseline, as an inpatient, and again at the end of study visit at 3 months (10-14 weeks).<br>- Data on readmissions at 1 year will be obtained from Hospital Episode Statistics. |

#### 4 TRIAL DESIGN

This is an open-label, single arm, single centre pilot study. The findings of this study, in terms of changes in self-reported alcohol intake, compliance and acceptability, will determine the feasibility of a randomized controlled trial to demonstrate clinical effectiveness.

#### 5 ELIGIBILITY CRITERIA

All patients with ALD **referred to** the Royal Free Hospital (RFH), **in either the inpatient or outpatient setting,** will be screened for eligibility for the study.

##### 5.1 Inclusion criteria

Inclusion criteria are:

- Subjects capable of giving informed consent
- Age 18-70
- ALD, including cirrhosis, up to Child-Pugh score of 7
- Possession of a smartphone
- Patients with alcohol dependence will be eligible for inclusion, providing inpatient detox has been successfully performed and there is no history of previous failed detox treatment.

##### 5.2 Exclusion criteria

Exclusion criteria are:

- Homeless or currently resident in a prison or rehab facility
- Inability to provide informed consent

### 6 TRIAL PROCEDURES

**Table 1 Schedule of Assessments**

| | Inpatient/ Outpatient | Outpatient | Outpatient |
|---|---|---|---|
| | Screening/Baseline | 3 months (10-14wks) | 14-52 weeks |
| Informed Consent | * | | |
| Demographics | * | | |
| Past medical history | * | | |
| Concomitant medication | * | | |
| QOL questionnaire SF36 | * | * | |
| Patient empowerment questionnaire (SUSTAINS) | * | * | |
| Dental examination | * | | |
| Stool Sample | * | * | |
| Saliva Sample | * | * | |
| Routine Biochemistry and Haematology blood samples (SOC) | * | * | |
| Adverse Events | * | * | |
| Provide device | * | | |
| Focus group assessment (up to 12 participants) | * | | * |

##### 6.1 Recruitment

Patients will be identified by the research team, or the alcohol team at Royal Free London[2] who review patients ALD. Screening **of inpatients and outpatients** according to the above criteria will then be performed by a member of the research team.

Patients who are pre screened but not recruited to the study will have anonymised data collected regarding age, gender, and the reason for non-eligibility or non-participation.

###### 6.1.1 Patient identification

Identification of participants will be performed either by the research team, or the alcohol liaison team at Royal Free London - part of the normal clinical team. These details will then be passed on to the research team who are also based at Royal Free London. Posters, leaflets or advertisements will not recruit participants. No personal information will be accessed by other patients or service users.

###### 6.1.2 Screening

Initial screening will be conducted by the research team or the alcohol liaison team. Specifically, screening will involve potential fulfillment of inclusion and exclusion criteria and establishing if the patient has a smart phone. No study interventions will be performed before consent.

##### 6.2 Consent

The Chief Investigator (CI) retains overall responsibility for the informed consent of participants at the Royal Free London site. All persons with responsibility for participation in the informed consent process will be duly trained and competent to participate according to the principles of Good Clinical Practice (GCP) and Declaration of Helsinki.

Participants are free to withdraw at any time from the trial without giving reasons and without prejudicing his/her further treatment. Participants will be provided with contact information for a member of the research team to obtain further information about the trial.

The consent process will involve:

- assessment of capacity
- discussion between the potential and an appropriate member of the research team regarding the nature and objectives of the study and possible risks associated with their participation
- the presentation of written material (PIS and consent document)
- the opportunity for potential participants to ask questions

Once the patient has signed the consent form a copy will be provided to the patient, the original retained in the study files and a copy retained in the medical notes. The subject will be provided with a sequential study number as follows;

ALC-001, ALC-002

---

[2] The alcohol service at the RFH is lead by Professor Kevin More (CI) and consists of specialists providing medical, social and psychological support for patients with alcohol problems.

###### *6.2.1 Additional consent provisions for collection and use of participant data and stool/saliva samples in ancillary studies:*

The stool and saliva samples obtained in this study will be stored and analysed for microbial analysis as stated in this protocol. Additionally, the stool and saliva samples will be stored for future, as yet unspecified, studies. Consent from participants will be sought for this aspect. No human genetic analysis will be conducted on these samples.

Participants will have the option of withdrawing from the storage of samples for ancillary research at any point, and samples will be destroyed.

6.2.2 Consent for attendance at focus group:

Study participants will be contacted by telephone, and asked for their willingness to participate in a focus group to explore the above issues. Two focus groups will be conducted, with four to six participants in each group. Each will last up to one hour and a half. If the individual agrees to take part, an information sheet and consent form will be provided and discussed before the focus group, with written consent being obtained before the focus group proceeds. The information sheet and consent form will clearly state that participants have the right to withdraw at any time without having to give a reason. Study procedures will be clearly explained to participants in terms of consent and anonymity, and their comprehension checked before the focus group commences to ensure informed consent.

##### 6.3 Baseline data

The baseline data set to be collected is:

- Age
- Gender
- Ethnicity
- Postcode
- Possession of smartphone and type of smartphone
- Previous 3 months of alcohol intake (by timeline follow back method)
- Quality of life questionnaire (SF36)
- Patient empowerment questionnaire (SUSTAINS)

Blood tests will be performed as SOC. The results of these tests will also be accessed by the research team and data collected on the following variables:

- Cabohydrate deficient transferrin
- Routine haematology and biochemistry

##### 6.4 Trial assessments

The specific trial procedures are:

- Blood tests at baseline and at 3 months (as part of SOC)
- Saliva and stool collection at baseline (as an inpatient or outpatient) and at 3 months. The exact timing of sample provision will be determine by the patient – can be anytime during the patient's initial inpatient stay/**outpatient visit**, or up to 6 weeks following discharge. The patient will be provided with collection pots to provide samples at any point around 3 months (10-14 weeks).
- Dental examination at baseline.
- The only study assessments will be at baseline (either as an inpatient or an outpatient) and at the 3-month (10-14 week) outpatient clinic visit, which is part of SOC. No dedicated study visits will be required.
- The patient will be supplied with the device and training provided. If the patient is having difficulty with the device, contact will be provided telephone or in person to provide further training as required.

##### 6.5 Long term follow-up assessments

There will be follow-up of participants beyond the 3-month study period. Data on hospital admissions up to 1 year will be assessed by accessing Hospital Episode Statistics data.

Patients that do not attend a follow-up clinic visit between 10-14 weeks will be contacted by telephone during the period 14-16 weeks following recruitment, to make arrangements to conduct the interview regarding alcohol consumption over the preceding 3 months (either in person or by telephone). If the patient cannot be contacted by week 16, then no further attempts to contact the patient by telephone will be made.

In case of premature discontinuation, similarly the research team will attempt to contact the patient by telephone within 14 days after discontinuation, to conduct the alcohol consumption interview. In this case, statistical analysis will be conducted by the Last Observation Carried Forward method.

Patients who complete the above assessments may be contacted up to 52 weeks following inclusion, to ask willingness to participate in a focus group (up to 12 participants, as described in 3.2 above).

##### 6.6 Withdrawal criteria

Patients may be withdrawn from study by the investigator for any of the following reasons:

- Compliance
- Patients who develop other health complications during the course of the study limiting their participation.
- Voluntary withdrawal from the study by patient or their legal representative.
- Investigator considers withdrawal from the study to be in patient's best interest.

##### 6.7 Storage and analysis of samples

Samples of faecal material and saliva will be collected from patients that have provided informed consent and have fulfilled the inclusion and exclusion criteria. All samples will be processed promptly to extract the bacterial material, and run on the sequencing machine in batches. Samples will be stored in the appropriate freezers until the end of the study after which they will be destroyed, except for those samples whereby subjects have consented for their samples to be used for unspecified future research use.

**Sample collection**

Faecal and saliva samples will be collected by clinical staff on the wards where the patient is admitted as an in-patient. The minimum amount of faecal material needed is two grams or a pea sized amount. The volume of saliva required would be a minimum of two milliliters. Universal containers will be used for sample collection. Two containers will be supplied per patient per visit, and labelled appropriately with the patients assigned study number.

Samples will be separated into smaller aliquots (approximately 0.2 g of faeces or 200µL of saliva) into sterile screw cap 1.5 mL tubes that have been appropriately labelled and then transferred to the Department of Microbial Diseases. The sample aliquots are then frozen at -80°C ± 10°C.

**Sample Analysis**

Samples will be processed and analysed in the Department of Microbial Diseases, University College London.

**Sample storage/destruction**

Once all of the sample analysis has been completed and all tests results have been obtained, all sample aliquots, original tubes, or bacterial extracts will be disposed of through autoclaving, and incineration. There will be no patient identifiable information on any of these samples.

**Storage**

In the study, saliva and faeces samples will be collected from patients in accordance with the patient consent form and patient information sheet and shall include all tissue samples or other biological materials and any derivatives, portions, progeny or improvements as well as all patient information and documentation supplied in relation to them. Samples will be processed, stored and disposed in accordance with all applicable legal and regulatory requirements, including the Human Tissue Act (HTA) 2004 and any amendments thereafter.

Samples should be aliquoted and stored at -80°C ± 10°C preferably on the same day of sample collection. Once the study is complete, samples will be destroyed except for those samples whereby subjects have consented for their samples to be used for unspecified future research use. Retained samples will be stored in a HTA licensed facility at the Eastman Dental Institute, UCL.

#### 7. Device

AlcoChange Smartphone Breathalyser

Manufacturer:

CyberLiver Ltd
First Floor Burnells
5 Garland Road
Stanmore
Middlesex
HA71NR
United Kingdom

Manufacturing Site:

ZANEC Soft Tech
Siri Towers, 2nd Floor,
No.3 & 4,Fourrts Avenue,
Okkiyam Thoraipakkam,
Chennai 600097, India

Means of Conformity:

Independent audit via BMQR against IVDD (98/79/EC)
Certificate No. CE 14780/R1 dated 26/03/2014
Class I *in vitro* diagnostic

Standards Used

EN 13612:2002, EN 13640:2002, EN ISO 18113-1:2011, EN 980:2008, and EN ISO 14971:2012

Fig 2. AlcoChange Device

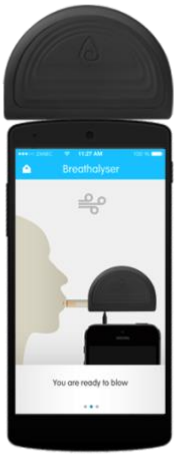

The device, developed by CyberLiver, is a combination of a smart phone app (AlcoChange mobile app) and battery-less breathalyser (AlcoChange Breathalyser) (Fig 2) and is classified as a Class I *in vitro* diagnostic CE Certified 20/04/14.

The AlcoChange app engages with its users continuously by nudging them to record what they have consumed. The app has highly personalised functions based on various parameters such as age, gender, preferred drinking days, preferred drinking times, and goals to achieve. Users can record their drink by choosing what they drank from the database of over 90,000 alcoholic beverages.

Fig 3.

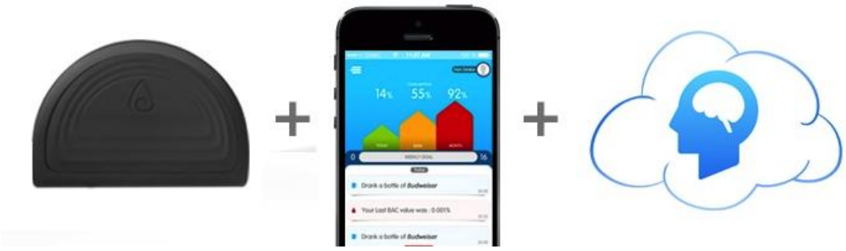

The app connects to a cloud based behaviour change platform (Fig 3). The platform collects data from all users and aggregates the same. It continuously analyses the users drinking patterns. The patent-pending behaviour change algorithms work to give personalized nudge messages to users to reinforce safe drinking patterns.

The AlcoChange smartphone breathalyser connects with the AlcoChange mobile app through a 3.5mm audio jack. Users can simply blow into the breathalyser to record their blood alcohol concentration. The AlcoChange breathalyser is battery less and uses a fuel cell sensor to determine the concentration of alcohol in breath.

The AlcoChange breathalyser requires calibration to retain the accuracy and it means that the device needs to be calibrated once a year. The AlcoChange Breathalyser is calibrated using advanced alcohol simulation equipment. The accuracy can fluctuate after 6 to 12 months of normal use depending on the number of tests performed & operating conditions. The product should be recalibrated at least every 6 to 12 months depending on usage.

Product Technical specifications:

| | |
|---|---|
| Operating voltage type | DC |
| Voltage Range | 1.8 - 4.5 V DC |
| Max Power Consumption | 3 MILLI WATTS |

| | |
|---|---|
| Dimension | 75mm X 49mm X 22mm |
| Weight in Grams | Less than 30 grams |
| Sensor Accuracy | +/- 0.01% BAC |

#### 8 Safety Reporting

##### 8.1 Definitions

**Used from MEDDEV guidance 2.7/3 and 2/12.1**

| Term | Definition |
|---|---|
| **Adverse Event** | Any untoward medical occurrence or effect in a patient treated on a trial protocol, which does not necessarily have a causal relationship with this treatment. An AE can therefore be any unfavourable and unintended sign (including an abnormal laboratory finding), symptom or disease temporally associated with the use of the trial treatment, whether or not related to the trial treatment. |
| **Abnormal Use** | Act or omission of an act by the OPERATOR or USER of a MEDICAL DEVICE as a result of conduct which is beyond any means of risk control by the MANUFACTURER. |
| **Corrective Action** | Action to eliminate the cause of a potential nonconformity or other undesirable situation. NOTE 1: There can be more than one cause for non-conformity.<br><br>NOTE 2: Corrective action is taken to prevent recurrence whereas preventive action is taken to prevent occurrence. |
| **Harm** | Physical injury or damage to the health of people, or damage to property or the environment. |
| **Incident** | "Any malfunction or deterioration in the characteristics and/or performance of a device, as well as any inadequacy in the labeling or the instructions for use which, directly or indirectly, might lead to or might have led to the death of a patient, or USER or of other persons or to a serious deterioration in their state of health.<br><br>Reference: Article 10 of the MDD<br><br>Note 1: There is a similar definition in Article 11 IVD Directive with minor wording differences. |
| **Serious Adverse Event** | A serious adverse event is any untoward medical occurrence that:<br>• results in death<br>• is life-threatening<br>• requires in-patient hospitalisation or prolongation of existing hospitalisation |

| | |
|---|---|
| | • results in persistent or significant disability/incapacity<br>• consists of a congenital anomaly or birth defect<br>• Other 'important medical events' may also be considered serious if they jeopardise the participant or require an intervention to prevent one of the above consequences<br>NOTE: The term "life-threatening" in the definition of "serious" refers to an event in which the participant was at risk of death at the time of the event; it does not refer to an event which hypothetically might have caused death if it were more severe. |

#### 8.2 Manufacturers role for Incident Reporting (CyberLiver)

The manufacturer (or their authorized representative) must submit an initial Incident report to the National Competent Authority (MHRA) for recording and evaluation. Each initial report must lead to a final report unless the initial and the final report are combined into one report. But not every INCIDENT report will lead to a corrective action.

#### 8.3 Recording and reporting of Incidents

Any event which meets all three basic reporting criteria A – C listed below is considered as an INCIDENT and must be reported to the relevant National Competent Authority (MHRA). The criteria are that:

**A: An event has occurred**

This also includes situations where testing performed on the device, examination of the information supplied with the device or any scientific information indicates some factor that could lead or has led to an event.

Typical events include, but are not limited to:

a) A malfunction or deterioration in the characteristics or performance.

A malfunction or deterioration should be understood as a failure of a device to perform in accordance with its INTENDED PURPOSE when used in accordance with the MANUFACTURER's instructions.

b) For IVDs where there is a risk that an erroneous result would either

(1) lead to a patient management decision resulting in an imminent life-threatening situation to the individual being tested, or to the individual's offspring, or

(2) cause death or severe disability to the individual or fetus being tested, or to the individual's offspring, all false positive or false negative test results shall be considered as events.

For all other IVDs, false positive or false negative results falling outside the declared performance of the test shall be considered as events.

c) Unanticipated adverse reaction or unanticipated side effect

d) Interactions with other substances or products

e) Degradation/destruction of the device (e.g. fire)

f) Inappropriate therapy

g) An inaccuracy in the labelling, instructions for use and/or promotional materials. Inaccuracies include omissions and deficiencies. Omissions do not include the absence of information that should generally be known by the intended USERs.

**B: The MANUFACTURER's device is suspected to be a contributory cause of the INCIDENT**

In assessing the link between the device and the INCIDENT the MANUFACTURER should take account of:

- the opinion, based on available evidence, of healthcare professionals;
- the results of the MANUFACTURER's own preliminary assessment of the INCIDENT;
- evidence of previous, similar INCIDENTs;
- other evidence held by the MANUFACTURER.

This judgement may be difficult when there are multiple devices and drugs involved. In complex situations, it should be assumed that the device may have caused or contributed to the INCIDENT and the MANUFACTURERs should err on the side of caution.

**C: The event led, or might have led, to one of the following outcomes:**

- death of a patient, USER or other person
- serious deterioration in state of health of a patient, USER or other person.

A serious deterioration in state of health can include (non exhaustive list):

a) life-threatening illness,

b) permanent impairment of a body function or permanent damage to a body structure,

c) a condition necessitating medical or surgical intervention to prevent a) or b). Examples: - clinically relevant increase in the duration of a surgical procedure, - a condition that requires hospitalisation or significant prolongation of existing hospitalisation.

d) any indirect harm (see definition under section 4.11) as a consequence of an incorrect diagnostic or IVD test result or as a consequence of the use of an IVF/ART device when used within MANUFACTURER´s instructions for use (use errors reportable under section 5.1.5.1 must also be considered).

e) foetal distress, foetal death or any congenital abnormality or birth defects.

**NOTE :** Not all INCIDENTs lead to death or serious deterioration in health. The non-occurrence of such a result might have been due to other fortunate circumstances or to the intervention of healthcare personnel.

It is sufficient that:

- an INCIDENT associated with a device happened, and
- the INCIDENT was such that, if it occurred again, it might lead to death or serious deterioration in health.

#### 8.4 Incident Reporting Timelines

**<u>Initial Report</u>**

**Serious public health threat:** IMMEDIATELY (without any delay that could not be justified) but not later than 2 calendar days after awareness by the MANUFACTURER of this threat.

**Death or UNANTICIPATED serious deterioration in state of health:** IMMEDIATELY (without any delay that could not be justified) after the MANUFACTURER established a link between the device and the event but not later than 10 elapsed calendar days following the date of awareness of the event.

**Others:** IMMEDIATELY (without any delay that could not be justified) after the MANUFACTURER established a link between the device and the event but not later than 30 elapsed calendar days following the date of awareness of the event.

In general, the report should be made to the National Competent Authority.

Final Report

The MANUFACTURER shall provide a follow-up-report to the National Competent Authority if the investigation time reaches the time line given to the National Competent Authority within the initial report.

**Note for this study reporting to MHRA will be undertaken by the chief Investigator or designee as required by the protocol and national regulations.**

##### 8.5 Recording and reporting of Adverse/Serious Adverse Events (non device related events)

The Investigator or appropriate site personnel will examine any subject experiencing an AE as soon as possible. The Investigator will do whatever is medically necessary for the safety and well-being of the subject. The subject will remain under observation at the Investigator‘s discretion. All AE‘s observed or reported following use of the AlcoChange device will be followed by the Investigator until resolved or until medically stabilized.

Whenever possible, the AE will be described on the appropriate CRF using standard medical terminology in order to avoid the use of vague, ambiguous, or colloquial expressions.
During the study, the Investigator or study site personnel will be responsible for querying and recording all adverse events (AEs) and serious adverse events (SAEs). Adverse events will be recorded from the date of signature of the informed consent until completion of the study at month 3 and will be evaluated by the Investigator for severity, seriousness, and relationship to study intervention, as outlined below. Untoward medical events which occur *after* signature of the informed consent *but before* use of the breathalyzer will be recorded as medical history. SAEs will be recorded from the signature of the consent form until 30 days after the last use of the breathalyzer.

Subjects will be asked at intervals throughout their assessments and again at the 3-month follow-up appointment about the occurrence of any adverse events. All AEs will be graded according to the criteria below. Additionally, the Investigator will determine the AE‘s relationship to the investigational device as described below. Adverse events will be followed by the Investigator until they have returned to baseline or stabilize.

**Serious Adverse Events: Reporting to Sponsor (UCL)**

Reportable Events

- any SAE
- any Investigational Medical Device Deficiency that might have led to a SAE if a) suitable action had not been taken or b) intervention had not been made or c) if circumstances had been less fortunate
- new findings/updates in relation to already reported events.

**Reporting Procedure**

Any SAE (or occurrence listed above under “Reportable Events”) that occurs during this study must be reported immediately (within 24 hours) to the sponsor’s office on the following email address:

Follow up information must be submitted promptly by faxing or e-mailing the updated SAE reporting forms (provided by UCL) to the sponsor office. The appropriate CRF(s) will be updated with any information collected while reviewing the SAE in a timely manner. The medical monitor is available to provide medical guidance regarding SAE reporting.

Serious Adverse Events:  Reporting to Ethics Committees
Any submission of a SAE that is unanticipated and related to the device must also be reported to the Ethics Committee (Health Research Authority).

It is the Investigators responsibility for reporting to the HRA.

Foreseeable Adverse Events
Given the underlying disease state for the study population for this trial, there are multiple expected serious adverse events.  For this trial, expected SAEs are presented below.

1) Anticipated Serious Adverse Events

- Variceal haemorrhage
- Spontaneous bacterial peritonitis
- Hepatorenal syndrome
- Multiple organ failure
- Death

- Disease progression
- Ascites
- Hepatic hydrothorax
- Non-lactobacillus bacteraemia
- Fungaemia
- Increased risk of infection
- Pre-renal failure
- Hepatic encephalopathy
- Peripheral neuropathy
- Autonomic neuropathy
- Hepatopulmonary syndrome
- Portopulmonary hypertension
- Cardiovascular insufficiency
- Hypotension
- Hepatocellular carcinoma (HCC)
- All non-HCC cancer

- Osteoporosis / Pathological fractures
- Hypoadrenalism
- Infertility

• Gynaecomastia• Hyperbilirubinaemia (jaundice)
• Elevated aminotransferases / Gamma-glutamyl transpeptidase
• Hypoalbuminaemia
• Abnormalities of serum sodium, potassium, calcium, magnesium, phosphate
• Anaemia
• Thrombocytopaenia
• Leucopaenia
• Coagulopathy
• Hyperammonaemia
• Hyperlactataemia
• Necessity for patients to undergo invasive procedures (e.g. transjugular liver biopsy and haemodynamic assessment)

The above, expected SAEs (except where the outcome leads to a patients death-see section 8.7), will not be subject to expedited reporting to the sponsor. These reports will be completed and provided to the sponsor in an agreed time frame.

2) Anticipated Non-serious Adverse Events

- Oedema
- Bruising
- Hair loss
- Amenorrhoea
- Hair loss
- Nausea
- Vomiting
- Constipation
- Diarrhea
- Headache

The above, expected AEs, will be recorded in the medical notes only.

Severity Grading for Adverse Events
The Investigator will assess all AEs according to the criteria described below. The severity of an event should be graded into one of three classes which describe the clinical severity of the event as it occurred.

The three classes of severity are:

- Mild / Grade 1: does not interfere with daily living and no medications are needed to relieve event;
- Moderate / Grade 2: somewhat interferes with daily living or medications needed to relieve event;
- Severe / Grade 3: incapacitating, activities of daily living are prohibited because of the event.

##### 8.6 Responsibilities

Principal Investigator (PI):

Checking for AEs when participants attend for treatment / follow-up.

1. Ensuring that all SAEs are recorded and reported to the Sponsor (UCL) within 24 hours of becoming aware of the event (except for those SAEs listed as expected and not requiring immediate reporting) and provide further follow-up information as soon as available. Ensuring that SAEs are chased with Sponsor if a record of receipt is not received within 2 working days of initial reporting.
2. Ensuring that AEs are recorded and reported to the Sponsor in line with the requirements of the protocol.
3. Reporting safety events to the HRA in line with national requirements.

Chief Investigator (CI) / delegate or independent clinical reviewer:

1. Clinical oversight of the safety of patients participating in the trial, including an ongoing review of the risk / benefit.
2. Using medical judgement in assigning seriousness, causality and expectedness of SAEs where it has not been possible to obtain local medical assessment.
3. Reporting any Incident to the MHRA as required by the protocol and regulatory requirements.

Sponsor (UCL):

1. Central data collection and verification of AEs and SAEs according to the trial protocol.
2. Reporting safety information to the CI, delegate or independent clinical reviewer for the ongoing assessment of the risk / benefit according to the Trial Monitoring Plan.

##### 8.7 Notification of deaths

- All deaths will be reported to the sponsor irrespective of whether the death is related to disease progression, the AlcoChange device, or an unrelated event.

#### 9 STATISTICS AND DATA ANALYSIS

The study has been independently reviewed by an external statistician, Yu Mei Ruby Chan and the results of the study will be analysed by both the CI and Statistician. As this is a pilot study no formal sample size calculation has been performed.

The following methodologies have been utilised:

- The primary outcome measure of self-reported alcohol consumption will be assessed at baseline and 3 months by TLFB. This will be assessed by paired t-test or Wilcoxon signed-rank test.

- Compliance with the AlcoChange device will be measured by quantifying frequency of interaction with the device. The relationship of compliance with the device and changes in alcohol consumption will be assessed by correlation and linear regression analysis.
- Secondary outcomes will include changes in microbial populations in saliva and stool, stratified by baseline dental health – these will be analysed by principal component analysis. Questionnaire data on HRQoL and patient empowerment will be analysed by Wilcoxon signed-rank test. Overall follow-up attendance rates at community alcohol services will be determined for study population, and compared with attendance rates during a corresponding period of time during the previous year.

### 10 DATA HANDLING

#### 10.1 Data collection tools and source document identification

##### Source Data

The source data to be accessed in this study will be patient demographic data, past medical history relating to alcohol use, blood tests as SOC, saliva and stool sample laboratory results, dental examination results. This data will be entered into the study specific Case Report Form (CRF) by the research team. Questionnaires (SF36 and SUSTAINS) will be considered source.

##### Case report forms

Case report forms (CRFs) will be used to record data from baseline and end of study (3-month) assessments as above. The CI will be responsible for keeping records of all participating patients (sufficient information to link records e.g., CRFs, hospital records and samples), all original signed informed consent forms and copies of the CRF pages.

##### AlcoChange Device Data

In this study, patients will enter personal data via the device, which will initially be stored on their own mobile phones. On availability of an internet connection, the data will be synchronised onto a centralised server hosted in a secure location which is NHS compliant.

#### 10.2 Data handling and record keeping

Patients will enter data on a routine basis during the study period, using the AlcoChange app that runs on Android or iOS operating systems. All data entered by the patients on their mobile phones will be transmitted to the 'CyberLiver' data platform through a web service that is securely made available only to the AlcoChange app over a secure HTTPS channel.

All patient identifiable data will be encrypted and stored in the database. The CyberLiver Platform database will be hosted on an NHS compliant environment.

Configuration and specification of the secure server is as follows;

Baseline Virtual Machines (VM) :2 x vCPU with 100Gb premium disk space (RAM, additional vCPU cores and extra disk space would be added as per requirement)

Failover & Recovery mechanisms will be put in place with high availability virtual machines having built in redundancy and automated restart capability of VM's on a new physical host within the cluster, in the event of a hardware or Hypervisor failure on the host.

Host-based backups would be performed nightly using the industry leading software. Backups of the VM container will be stored on-site unless otherwise specified, to a 14 point retention period; backup disk space up to 50% of allocated disk.

A Virtual Machine would be used which would ensure that dedicated physical disks assigned to virtual database machines ensuring disk IO performance matches or betters that found on physical machine databases.

The servers will be secure and segregated from all other client databases and/or applications within a dedicated VLAN running under a dedicated operating system.

High availability recovery will be in place within minutes in the event of critical failures.

Server patching, updates, application and database administration will all be managed by the CyberLiver team.

Token based secure 2-Factor Authentication will be in place in addition with N3 connectivity.

.

The platform will also have a secure web-based portal that will be accessible only by the alcohol liaison team and the research team. Unique sets of credentials for each user requiring access will be provided. All users will only have read-only access to patient data. All user actions will be monitored with audit logs of every action performed by the users.

#### 10.3 Data transfer (handling, processing and storage)

In the study, basic demographics, name, and alcohol history together with drinking history will be collected from patients in accordance with the patient consent form, patient information sheet and sections 6.3, 6.4 and 6.5 of this protocol.

The basic demographics, name, and alcohol history together with drinking history will be appropriately sent to PHARMExcel Ltd for study monitoring and to Dr Yu-Mei Ruby Chang for statistical analysis. Professor Moore and Dr Mehta will act as the data controllers of such data for the study.

Professor Moore and Dr Mehta will process, store and dispose of basic demographics, name, and alcohol history together with drinking history, in accordance with all applicable legal and regulatory requirements, including the Data Protection Act 1998 and any amendments thereto.

##### 10.4 Access to Data

Direct access will be granted to authorised representatives from the Sponsor, host institution and the regulatory authorities to permit trial-related monitoring, audits and inspections.

##### 10.5 Archiving

UCL and each participating site recognise that there is an obligation to archive study-related documents at the end of the study (as such end is defined within this protocol). The Chief Investigator confirms that he/she will archive the study master file at Royal Free Campus, UCL for the period stipulated in the protocol and in line with all relevant legal and statutory requirements. The Principal Investigator at each participating site agrees to archive his/her respective site's study documents for 5 years and in line with all relevant legal and statutory requirements.

#### 11 MONITORING, AUDIT & INSPECTION

Since this is a low-risk trial, monitoring will be limited to a single data monitoring visit, by a contract research organisation (Pharmexcel Ltd) prior to the completion of the study.

#### 12. INSURANCE

University College London (UCL) holds insurance against claims from participants for injury caused by their participation in the trial. Participants may be able to claim compensation if they can prove that UCL has been negligent. However, as this trial is being carried out in a hospital, the hospital continues to have a duty of care to the participant of the trial. University College London does not accept liability for any breach in the hospital's duty of care, or any negligence on the part of hospital employees. This applies whether the hospital is an NHS Trust or otherwise.

Participants may also be able to claim compensation for injury caused by participation in this trial without the need to prove negligence on the part of University College London or another party. Participants who sustain injury and wish to make a claim for compensation should do so in writing in the first instance to the Chief Investigator, who will pass the claim to the Sponsor's Insurers, via the Sponsor's office.

Hospitals selected to participate in this trial shall provide negligence insurance cover for harm caused by their employees and a copy of the relevant insurance policy or summary shall be provided to University College London, upon request.

The manufacturer, CyberLiver, holds indemnity to cover the malfunction and breakdown of the device.

#### 13. APPENDICIES

##### 13.1 Appendix 6 – Amendment History

| Amendment No. | Protocol version no. | Date issued | Author(s) of changes | Details of changes made |
|---|---|---|---|---|

Topic Guide: AlcoChange Study (IRAS: 190682).
Authors: Aida Nadar, Lorraine McDonagh

Thank you so much for your time and for coming along to talk with us today. My name is Aida and I am a research intern here at the royal free.– The aim today is to get a deeper understanding of your experience with AlcoChange and finding out what we can change to make the app more useful for you (and other people like yourself in the future)

- Go through information sheet
- Go through consent form – have them sign it

The plan for this afternoon is to firstly discuss your experience with alcohol and ARLD and the steps you have taken in the past to stop drinking.

Following this we will discuss your experience with the app and what potential improvements could be made to it in the future.

Does that all sound ok?

Before we start it is important for you to know that there are no right or wrong answers. We are just exploring this topic and are really interested in hearing about your opinions. If you don't feel comfortable answering any questions, you don't have to.

This is just a relaxed discussion so respect opinions and let everyone have their say. Listen / talk to each other not me. Everything discussed here today will be confidential.

Do you have any questions before we start? This session will be recorded is that ok - all names will be anonymized.

Turn on recorder.

Have people introduce themselves (use name tags)

1. Why did you decide to take part in this study?

2. Can you tell me a bit about your usual drinking patterns?
    - ↳ Potential probes:
        - ↳How long have you experienced this…?
        - ↳How serious would you say this is for you?
        - ↳Does this have an impact on your day to day life?
        - ↳When were you first diagnosed with ARLD

3. What were your motivations for taking part in the study?

4. What prompted/motivated you to want to achieve abstinence?
   (Ask about context of experiences)

5. What steps have you taken in the past to stop drinking?
   - ↳ Prompt: which did you find most useful – what was your longest period of abstinence achieved?
   - ↳ What triggers you to resume drinking?

- Were you engaging with any other services or using any other tools whilst on the AlcoChange?

**Prompt: If you have used other alcohol services or face to face support groups – how do you think AlcoChange compares in terms of how effective it was?**

**QUESTIONS RELATED DIRECTLY TO THE APP**

6. Overall, what do you think of the app – has it been helpful / unhelpful?

7. How often did you use it?
   - ↳ Probe: did certain events or times make you use it more

8. How long did it take for you to learn how to use this application?
   - ↳ Prompt. If necessary, – What kind of work and skills are needed to use the application well?

9. What benefits, if any, did you get from using the app?

10. Do you think this app has any drawbacks – or aspects you found difficult?

11. Have you ever used an app to manage your drinking before?
    - ↳ Prompt: if so how do they compare

12. Would you recommend the app to a friend?

**FEATURES**

13. Which features did you dislike
    - ↳ Prompt: did you use these the least?

14. Which features did you particularly like?
15. How user-friendly would you say it is?

16. Was there anything that made you unsure about using the app?

**SPECIFIC FEATURES [Now let's chat about some of the specific features of the app…]**

17. What were your thoughts on the breathalyzer feature?
    - ↳ Did you use it regularly
    - ↳ Did you find it helpful/not helpful etc…

18. Did you ever contact the helpline or a friend through the app –
    - ↳ Prompt: If so, how was that?

19. Did you log your appointments on the app calendar

20. Did you regularly log cravings? / → if so did this help you overcome them

21. Did you log when you had a drink?

**POTENTIAL IMPROVEMENTS** [Now let's chat about potential improvements to…]

22. What are your thoughts on how we could improve AlcoChange

23. Are there any features you would like to see added?
    - ↳ Probe: Can you tell me more about that

**BEHAVIOUR CHANGE:**

1. What changes in your behaviour did you have to make while using the application?
    - ↳ Prompt: If necessary, how has use of this application influenced your drinking habits.
    - ↳ Prompt: Did any specific features help bring this changes about….

2. How easy was it to adopt these changes?
    - ↳ Prompt: What other support did you draw on, in addition to this app?

3. Do you feel you have learned anything from using Alcochange?
    - ↳ Probe: Can you tell me more about that?

*MAY ALSO WANT TO ASK, what impact (if any) has it had on their life –*

4. Has using Alcochange had any impact on your…
    i. work life
    ii. social life and relationships (family, partners, friends)
    iii. (perceived) psychological impact (mental health and well-being)
    iv. physical health etc

↳ Probe: Can you tell me more about that?

↳ Probe: What exactly was that impact?

↳ Prompt: How do you think has Alcochange helped achieve this impact

↳ Prompt: Do you think this impact was short-lived or will it be sustained

**CLOSING**

24. Do you have any final thoughts or comments on how we could improve AlcoChange and the way that it is offered to patients?

25. Any further comments or anything else you would like to add to the discussion?

**Thank you very much for coming**

**Calibration of the AlcoChange breathalyser:**

The AlcoChange breathayser is designed as a battery-less device, which is powered by, and operated through, the smartphone. The attachment to the smartphone is through the 3.5mm connection. Data is securely stored in a cloud platform, and idenitity of the user is veriified through the front smartphone camera before breath alcohol measurements are stored. This feature validates the identity of the individual being tested.

Alcohol content is measured using fuel cell sensors. Calibration is required annually.

Example calibration data is provided below. Six independent alcohol measures were taken in three indivduals, at 20 minutes after consumption of 24-32g ethanol. Alcohol was measured by blood content, breath alcohol (BACtrack beathalyser) or AlcoChange breathalyser (Figure S1).

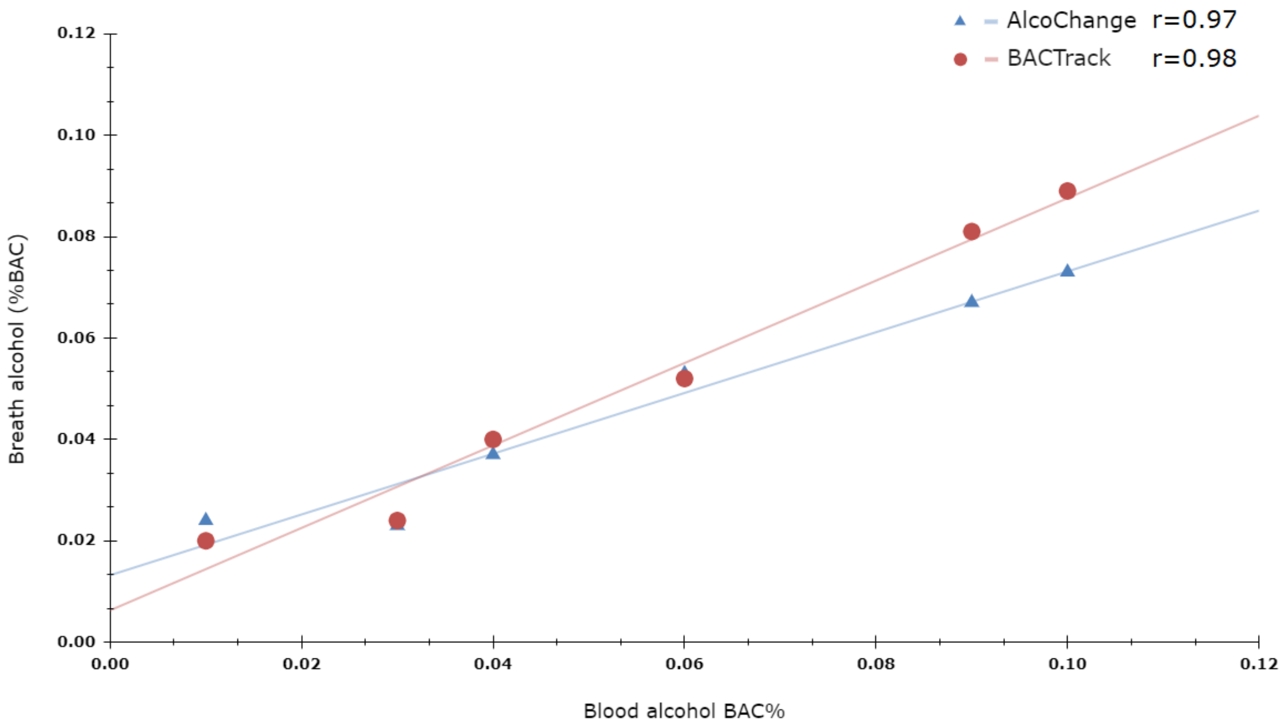

**Figure S1: Calibration of AlcoChange breathalyser against blood alcohol content and BACTrack breathalyser measurement.** n=6 observations, Pearson's correlation performed.
